# Characterizing Public Sentiments and Drug Interactions during COVID-19: A Pretrained Language Model and Network Analysis of Social Media Discourse

**DOI:** 10.1101/2024.06.06.24308537

**Authors:** Wanxin Li, Yining Hua, Peilin Zhou, Li Zhou, Xin Xu, Jie Yang

## Abstract

**Objective:** Harnessing drug-related data posted on social media in real time can offer insights into how the pandemic impacts drug use and monitor misinformation. This study developed a natural language processing (NLP) pipeline tailored for the analysis of social media discourse on COVID-19 related drugs.

**Methods:** This study constructed a full pipeline for COVID-19 related drug tweet analysis, utilizing pre-trained language model-based NLP techniques as the backbone. This pipeline is architecturally composed of four core modules: named entity recognition (NER) and normalization to identify medical entities from relevant tweets and standardize them to uniform medication names, target sentiment analysis (TSA) to reveal sentiment polarities associated with the entities, topic modeling to understand underlying themes discussed by the population, and drug network analysis to potential adverse drug reactions (ADR) and drug-drug interactions (DDI). The pipeline was deployed to analyze tweets related to COVID-19 and drug therapies between February 1, 2020, and April 30, 2022.

**Results:** From a dataset comprising 2,124,757 relevant tweets sourced from 1,800,372 unique users, our NER model identified the top five most-discussed drugs: Ivermectin, Hydroxychloroquine, Remdesivir, Zinc, and Vitamin D. Sentiment and topic analysis revealed that public perception was predominantly shaped by celebrity endorsements, media hotspots, and governmental directives rather than empirical evidence of drug efficacy. Co-occurrence matrices and complex network analysis further identified emerging patterns of DDI and ADR that could be critical for public health surveillance like better safeguarding public safety in medicines use.

**Conclusion:** This study evidences that an NLP-based pipeline can be a robust tool for large-scale public health monitoring and can offer valuable supplementary data for traditional epidemiological studies concerning DDI and ADR. The framework presented here aspires to serve as a cornerstone for future social media-based public health analytics.

## 1. Introduction

The emergence of the COVID-19 pandemic has induced an immediate need for effective pharmacotherapies. While the development and application of such therapies are critically important, they are also influenced by an array of political, economic, and social factors. For example, public pronouncements by high-profile figures, such as former U.S. President Donald Trump’s endorsement of hydroxychloroquine, have led to its irrational use and consequential public health crises.[1] Traditional pharmacovigilance mechanisms, reliant on clinical trials and formal reporting systems like MedWatch and DrugBank,[2–4] offer valuable but lagged information. These traditional approaches are plagued by inefficiencies, reporting biases, and a lack of timeliness, thereby lacking comprehensive coverage of the population’s sentiments and experiences.[5–8]

In this context, real-time public comments on pharmacotherapies such as medications on social media provide a valuable resource for complementing research on drug use or repositioning for COVID-19. In addition to the fast accessibility, timeliness, and comprehensive population coverage, social media can also supply real-world evidence on how people respond to different drugs, thus helping researchers mine novel drug potency or side effects.[9–11] Social media also offer data on drugs not typically included in pharmacovigilance datasets, such as over-the-counter drugs,[12] herbal remedies,[13] and other non-traditional treatments.[14] However, the sheer volume and noise in social media data require robust computational methodologies for effective analysis.[15]

Natural language processing (NLP) technologies offer a solution to these challenges. Earlier studies, such as the study conducted by Aramaki et al. in 2011, demonstrated that Twitter data could be mined to monitor influenza outbreaks using machine learning and rudimentary NLP techniques.[16] Contemporary research in this domain has benefitted immensely from advancements in deep learning and specialized NLP tools, such more specialized NLP tools for analyzing social media data[17] and public health datasets for social media posts.[18] These have made it increasingly feasible to sift through large volumes of colloquial, noisy text to extract meaningful insights on public health.

Within the realm of COVID-19 drug research, substantial efforts have been made to analyze pharmacotherapy-related topics during COVID-19. For example, Hua et al. utilized BERT models to examine public perceptions of specific, albeit controversial, medications and found these perceptions to be heavily skewed by misinformation and partisan biases.[19] However, the study suffered from methodological limitations, including a narrow and subjectively chosen selection of drugs (i.e., hydroxychloroquine and ivermectin versus Molnupiravir and Remdesivir). Similarly, Wu et al. made the very first attempts to construct co-occurrence networks to study symptomatology, but their technique was solely based on lexicon matching, thus subject to false positives.[20] There is still a lack of data-driven pipeline with state-of-the-art NLP tools and other big data analysis techniques to automatic extract drug information through social media data from the view of public health.

To address these gaps, this study employs NLP methodologies and network analysis for an extensive assessment of COVID-19 drug-related discourse on social media. We contribute to the existing literature in several ways:

1. Employing deep learning methodologies for named entity recognition (NER), thereby reducing the false positives associated with traditional keyword matching.
2. Re-examining public sentiments and concerns regarding COVID-19 medications, utilizing target sentiment analysis (TSA) and topic modeling.
3. Conducting a comprehensive assessment of adverse drug reactions (ADR) and drug-drug interactions (DDI) through network analysis techniques.

We demonstrate that our integrated NLP pipeline can serve as a robust framework for extracting and analyzing drug-related information, thereby enhancing the scope and effectiveness of social media-based pharmacotherapy analysis.

## 2. Methods

As shown in Fig. 1, the study workflow is organized in three primary stages: data collection, development of an NLP pipeline, and subsequent data analysis using the constructed pipeline. Initially, we curated a dataset of English tweets related to COVID-19. After a preprocessing phase that excluded tweets with URLs, a NLP pipeline was developed to extract and normalize the durgs/diseases mentioned in these tweets. Finally, we examined the time trends of drug mentions, public sentiment, and discussion topics towards drugs, as well as the co-occurrence network of drug-drug and drug-disease pairs. Ethical approval for this study was granted by the Institutional Review Board of Zhejiang University.

**Fig. 1.**
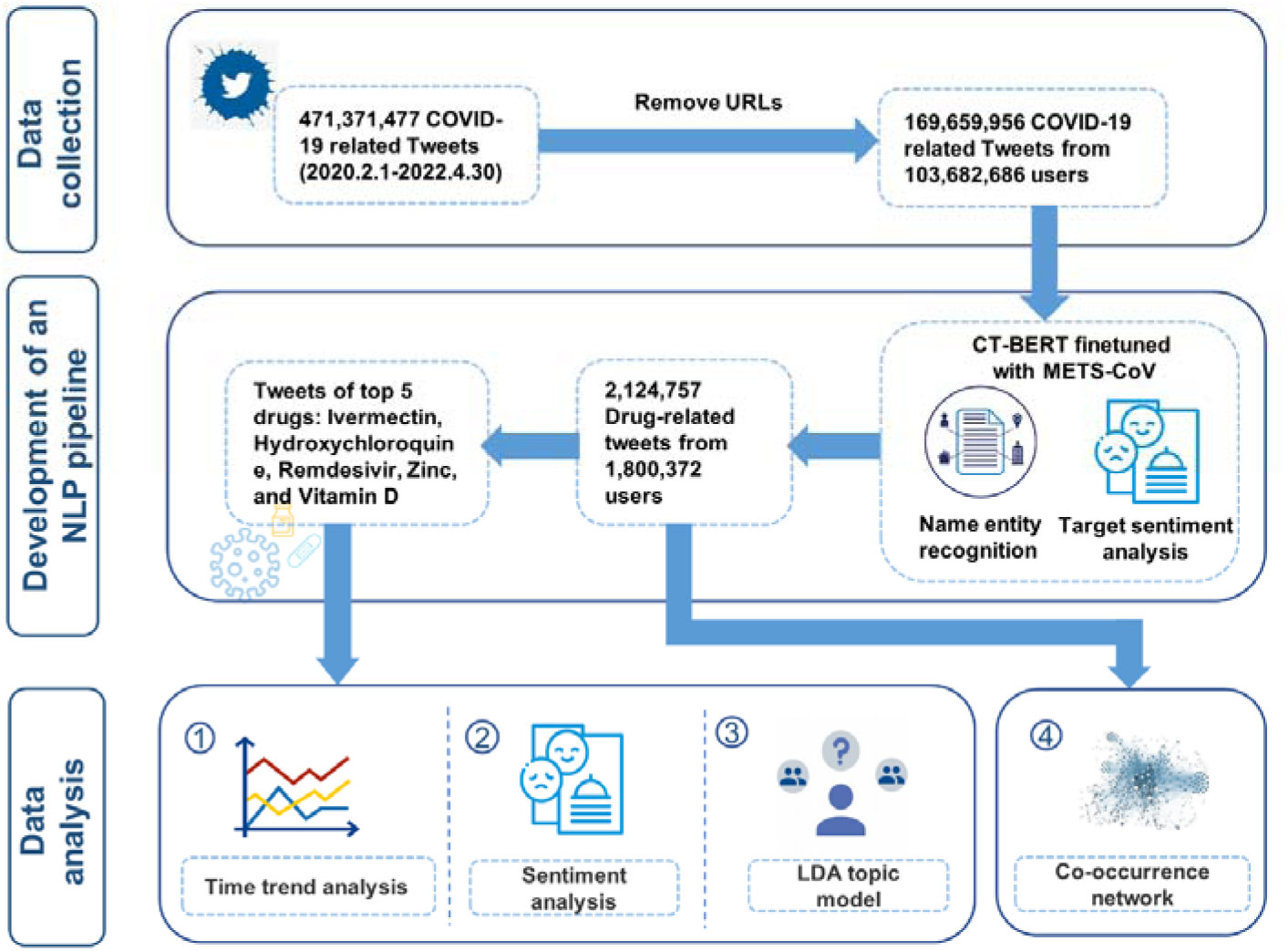
Workflow of drug analysis with NLP on Twitter. NLP: natural language process; CT-BERT: COVID-Twitter-BERT; METS-CoV: Medical Entities and Targeted Sentiments on COVID-19-related tweets, NER dataset containing medical entities and targeted sentiments from COVID-19 related tweets.[18]

### 2.1 Data collection and preprocessing

The used tweets were obtained from a public dataset provided by Chen et al[21] and were downloaded using Twitter’s API. The downloaded data included full tweet texts and corresponding metadata such as timestamps and user information. Tweets containing URLs were excluded from the analysis, as they often only contained summaries or quotations of the original tweet. The data collection process adhered to Twitter’s privacy and data use management policies.

### 2.2 NLP pipeline development

The NLP pipeline consists of four principal modules: Named Entity Recognition (NER), Target Sentiment Analysis (TSA), topic modeling, and drug network analysis. For the NER and TSA modules, we leveraged state-of-the-art models developed in our prior work Medical Entity and Targeted Sentiment on COVID-19 Related Tweets (METS-CoV).[18] Details on model construction can be found in Fig. A.1.

#### 2.2.1 Named entity recognition (NER) and normalization

The NER model aims to extract drug entities from tweets. The model we developed, CT-BERT-NER,[18] was constructed using the COVID-Twitter-BERT (CT-BERT) framework, a widely adopted language model pre-trained on 160 million COVID-19-related tweets. It was trained on the entire training set of the NER subset of METS-CoV.[18] Upon evaluation, it showed F1 scores of 86.35% for drug entity recognition and 81.85% for symptom entity recognition on the corresponding test set.[18] We used the model trained on all entity types instead of on drug entities only to enable the nuanced differentiation of drug entities from other types of entities.

To standardize colloquial expressions of drugs among the extracted entities, we manually searched from Wikipedia for NER-identified drug entities with frequency more than 1000 to map colloquial drug expressions and their standardized concepts (i.e., drug trade names, chemical names, and generic names). We conducted an accuracy assessment using a random sample of 50 tweets for each of the top five most frequently mentioned drugs and symptoms, as identified through two methods: NER combined with lexicon-based extraction (NER + lexicon) and lexicon-based extraction alone. Our results demonstrated that the NER + lexicon method achieved an accuracy rate of 100%, significantly surpassing the 89% accuracy achieved by the lexicon-only approach. Further details on this comparison are available in Table A.1.

#### 2.2.2 Targeted sentiment analysis (TSA)

The TSA module aims to quantify user’s sentiment toward specific drug entities within tweets. The TSA model, CT-BERT-TSA[18], is a three-class model developed based on the BERT-SPC framework.[22] Similar to BERT-SPC, CT-BERT-TSA model treats targeted sentiment analysis as a sentence pair classification task, by appending the identified drug entity to the end of the tweet context and then feeding this sentence pair into CT-BERT model for three-class prediction (i.e., positive, neutral, and negative). Upon testing on the TSA test set of METS-CoV, the model showed an F1 score of 62.67% and an accuracy rate of 75.07%,[18] across four entity types — Person, Drug, Disease, and Vaccine.

#### 2.2.3 Topic model analysis

To discern prevailing public interests in the most discussed drugs, we implemented Latent Dirichlet Allocation (LDA) for topic modeling, utilizing the *LdaModel* function from the *Gensim* package.[23] Topic numbers were determined based on conventional evaluation metrics, including low perplexity and high coherence scores.[24], [25] Detailed methodologies are delineated in Fig. A.2.

#### 2.2.4 Drug network analysis

For elucidating potential relationships among drugs, we constructed this drug network analysis module to generate incidence matrices and visualize co-occurrence networks using Gephi.[26] For enhanced comprehensiveness, we incorporated a variant supported by the Anatomical Therapeutic Chemical Classification System (ATC),[27] in addition to the Gephi-based visualization. In addition, we used the NER model to extract symptom entities and normalize them through a pre-summarized lexicon list[28] to extend our analysis to drug-symptom networks. The constructed networks feature nodes representing either frequently occurring drugs or symptoms. As our focus is not on causal relationships but rather on the interplay between entities, we employed undirected graphs and used semantic cosine similarity[29] as the distance metric.

### 2.3 Pipeline deployment

Upon completion of the NLP pipeline, we proceeded to its deployment on the pre-processed dataset of COVID-19-related tweets. We first applied the NER and normalization module on the pre-processed dataset to extract and standardize drug entities to drug concepts. Following this standardization, we conducted a distributional analysis of drug mentions to discern time trends, thereby capturing the evolving popularity of these drugs. We also gather related news and the trend of weekly new COVID-19 cases to show a more holistic view of the shift of drug popularity over time. For clarity and simplicity, we only illustrate the top five most discussed drugs.

Subsequently, we used the TSA model to assign each drug entity of the five types a sentiment type. To gain a deeper understanding, we also conducted a time-trend analysis on the positive and negative tweets for the five drugs and visualized the results. Building upon our understanding of public sentiment, we turned to topic modeling via LDA to explore the thematic concentrations in the discourse surrounding these drugs. The model yielded the 20 most probable keywords and bigrams for each identified topic, enabling us to summarize the primary themes. We further analyzed the topic distribution associated with each of the top five drugs.

Finally, we constructed co-occurrence networks for drug-drug and drug-symptom interactions to provide a relational overview that complements our earlier analyses. All drugs with more than 1,000 mentions over time were included in the analysis. Meanwhile we also zoomed in to analyze the five most-discussed drugs.

## 3. Results

### 3.1 Data summary and trends of drug mention tweets

This study used a dataset consisting of 471,371,477 COVID-19-related tweets in English, which were collected between February 1, 2020 and April 30, 2022. After excluding tweets containing URLs, the final dataset used for this study consisted of 169,659,956 tweets from 103,682,686 user. Using CT-BERT-NER, we identified 2,124,757 drug-related tweets from 1,800,372 unique Twitter users, accounting for approximately 1.25% of the raw COVID-19-related tweets dataset. Table A.2(in supplementary materials) provides more detailed statistical results of the medical entity recognition.

Table A.3 presents the 67 most frequently mentioned drugs, each with an occurrence exceeding 1000 times. The most frequent taxonomies are Anatomical Therapeutic Chemical Classification System (ATC)[27] is N (nervous system drugs) and J (anti-infective drug). We ranked the total occurrence of all drugs and identified the top five most-mentioned drugs: Ivermectin, Hydroxychloroquine, Remdesivir, Zinc, and Vitamin D to visualize their weekly time trends. Fig. 2 presents these temporal trends.

**Fig. 2.**
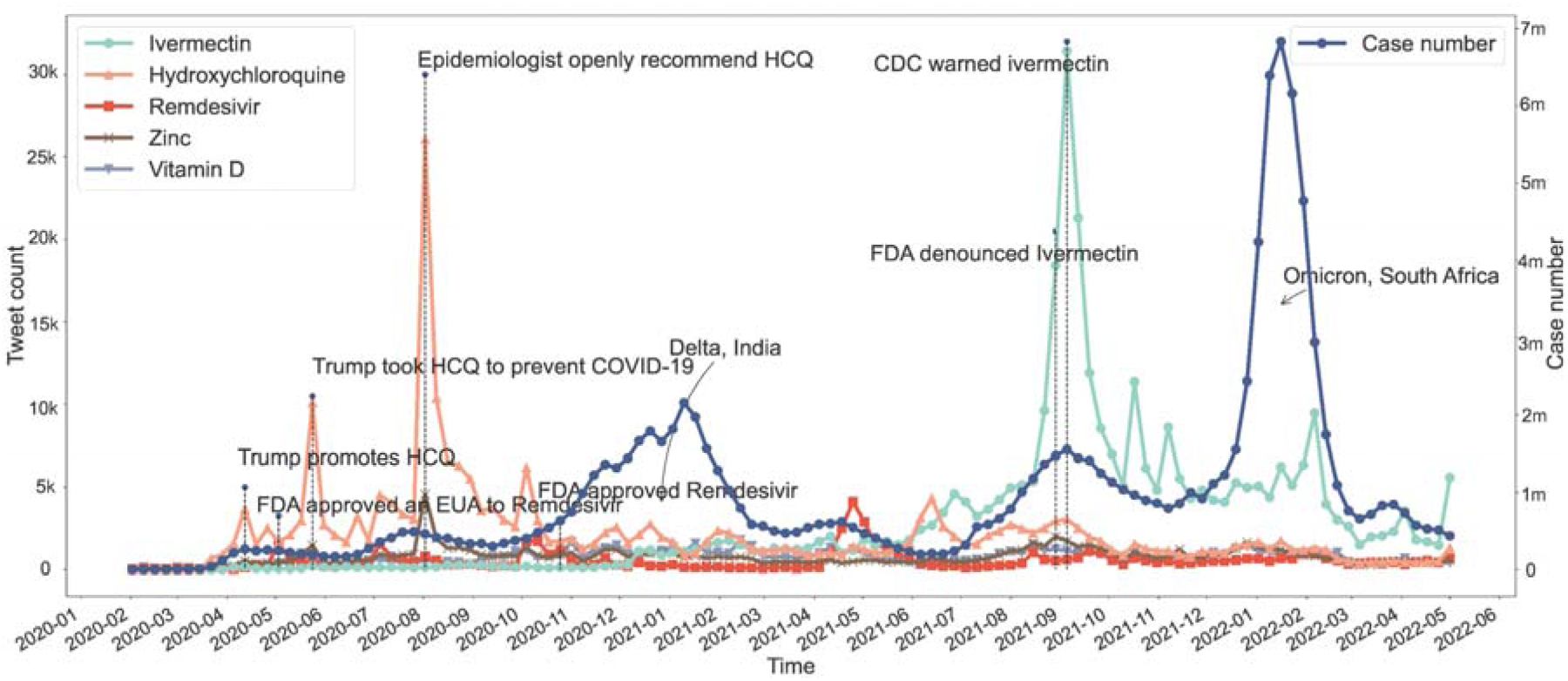
Weekly popularity trends of the top five most-mentioned drugs on Twitter examined with COVID-19-related tweets collected between February 1, 2020 and April 30, 2022. The left Y-axis represents the total number of tweets for each drug in a given week (unit: thousand tweets). The right Y-axis represents weekly new case count (unit: million tweets). The new case counts were collected from World Health Organization (WHO)[30] on a weekly basis, beginning on February 1st, 2020. Given that the dataset is confined to English-language tweets, the scope of new case counts was likewise restricted to the top four English-speaking nations with the highest Twitter activity: the United States, the United Kingdom, the Philippines, and Canada.[31]

Among the five drugs, the public focused mostly on repurposed drugs (i.e., hydroxychloroquine and ivermectin), followed by daily supplements (i.e., zinc and vitamin D). The only officially approved drug among the five, Remdesivir, received the least attention. The frequency of discussion of hydroxychloroquine and ivermectin fluctuated significantly across time, which seemed to be related to relevant news events or policies (marked in Fig.2). In the early stage of the pandemic, drug-related discussions focused on hydroxychloroquine, with two prominent peaks occurring at May 24^th^ and August 2^nd^ of 2020. Discussion of ivermectin began to increase in the later stages of the pandemic, with only one prominent peak located at September 5^th^ of 2021. In contrast, Remdesivir received the least public attention, which increased only sporadically throughout the pandemic, with no apparent pattern and a much lower peak on May 3^rd^ of 2020. As supplements to COVID-19 treatments, vitamin D and zinc elicited much less public interest than ivermectin and hydroxychloroquine, with no significant outbreaks or visible patterns.

### 3.2 Changes in sentiment for five most frequent mentioned drugs

We calculated sentiment proportion for the five drugs and the weekly time trends of positive and negative tweets. Fig. 3a shows the visualization of the overall attitude proportions. The public tended to hold positive and neutral attitudes toward the repurposed drugs, ivermectin and hydroxychloroquine. The immune supplements, zinc and vitamin D, were frequently mentioned with positive sentiments. The only COVID-19 drug approved by the FDA, Remdesivir, received the lowest positive attitude (12.8%), far lower than those of the other drugs.

**Fig. 3.**
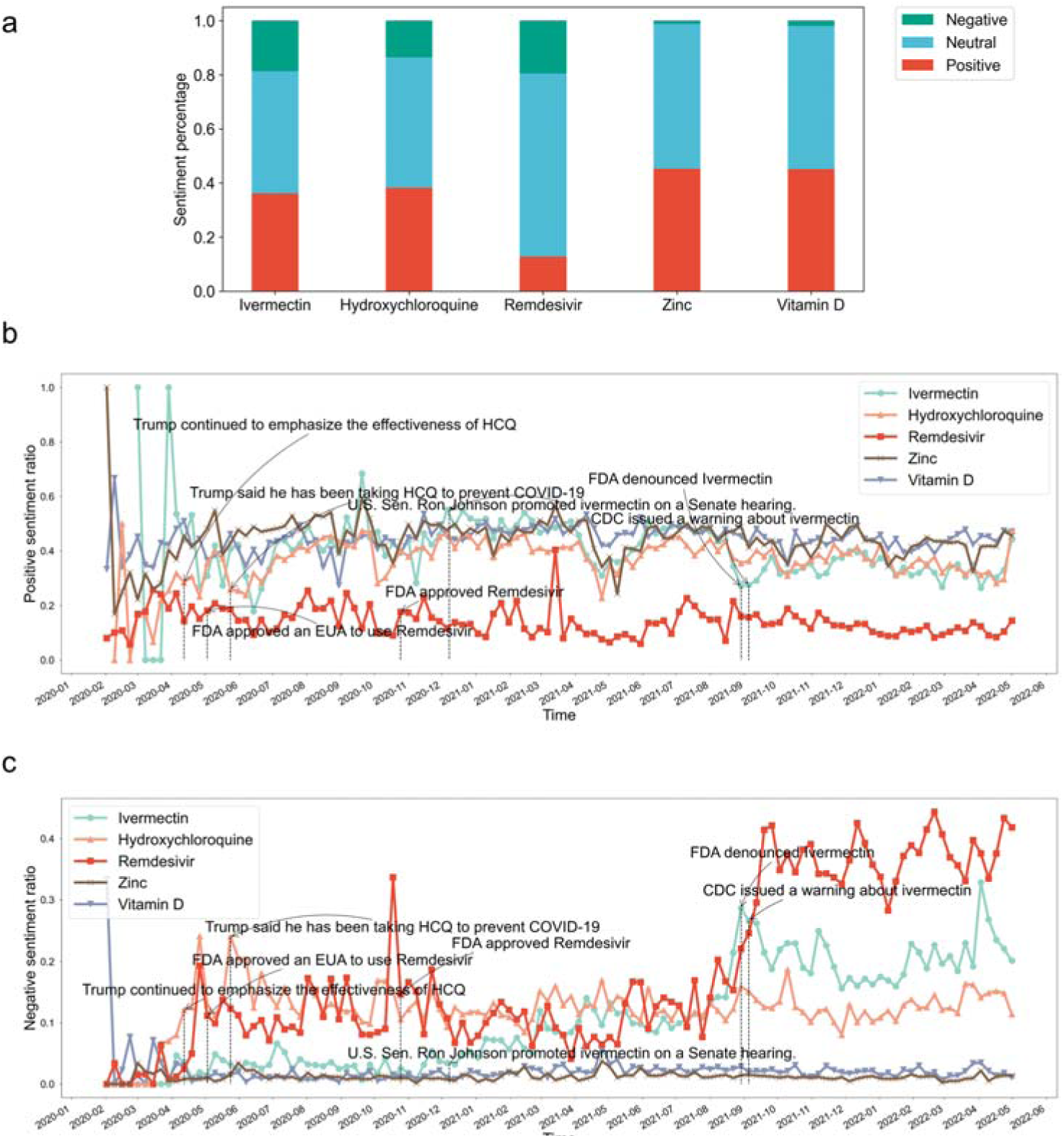
Sentiment analyses of the five top-discussed drug from February 1, 2020 to April 30, 2022, grouped according to their polarity, including (a) sentiment distribution, (b) weekly ratio of positive tweets, and (c) weekly ratio of negative tweets. The denominator of the percentage was the entities with sentiment.

Fig. 3b and 3c presents weekly trends of tweets expressing positive and negative attitudes, respectively. The major turning points of the trends tend to coincide with new government policies, major social events, and research findings. The criticism of remdesivir (Fig. 3c) and ivermectin increased over time since September 2021, and the turning point for remdesivir came at almost the same time of emerging studies showing that the drug is ineffective[32] and has severe side effects.[33–35] For ivermectin, public sentiment was associated with announcements of health authorities and celebrity effects. For example, the FDA denouncing the use of ivermectin for COVID-19 on August 29^th^, 2021 had simultaneously increasing negative discussions.

### 3.3 Topics distributions of drug mentioned tweets

We applied the LDA topic model to all drug-related tweets and obtained 15 general topics based on their relatively high topic coherence scores and low confusion levels (further discussed in Fig. A.3). We display the corresponding top 20 most likely keywords in Table 1, and assigned a theme for each topic from these keywords. The topic "clinical treatment effect of drugs" dominated the discussions, accounting for 13.60% of all related tweets. Other popular topics included "physical symptoms" (11.84%) and "causes of death" (9.28%), closely followed by topics such as "immune response" (8.14%), "general treatment" (8.23%), and "daily supplement intake" (7.27%).

**Table 1.**
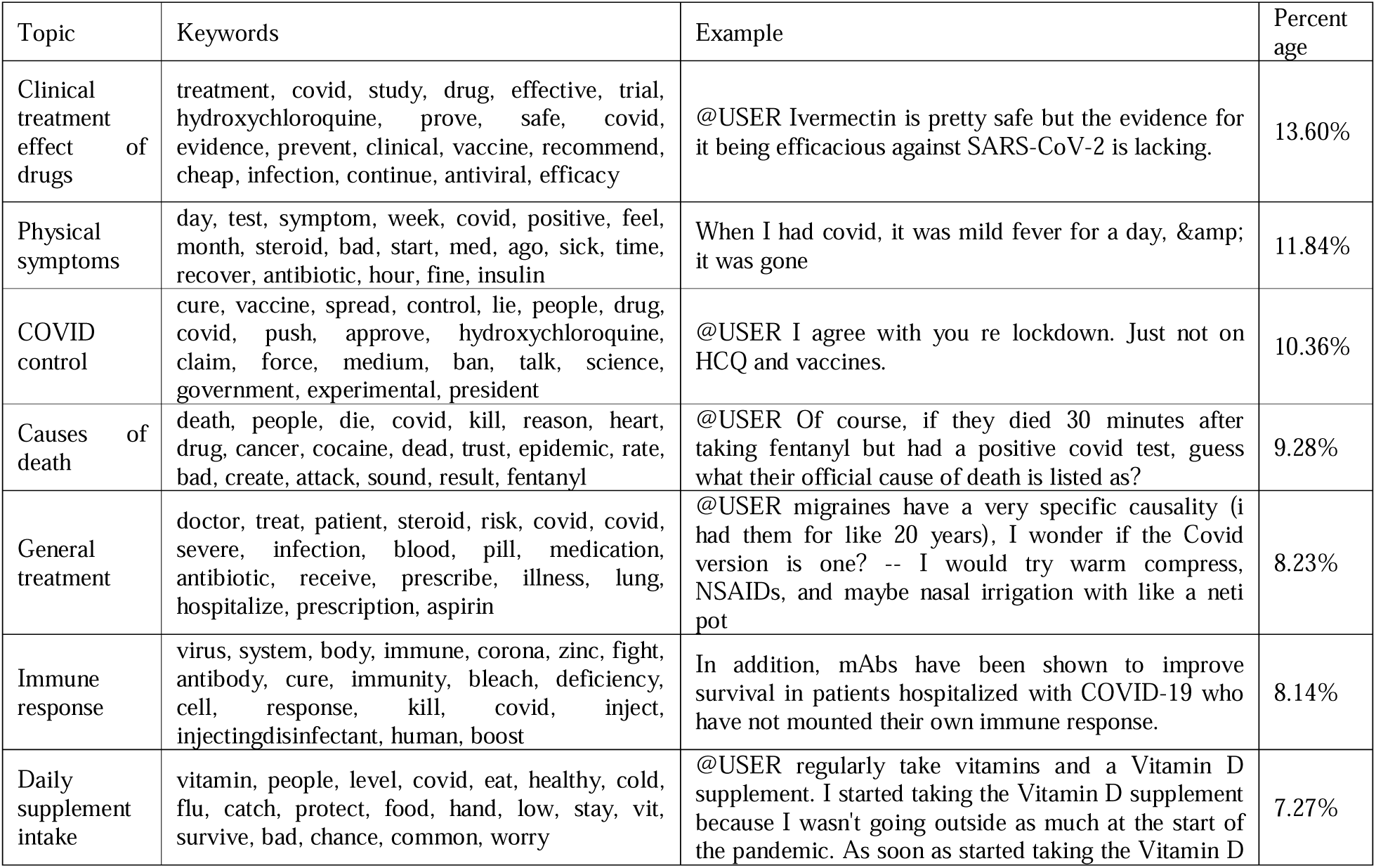
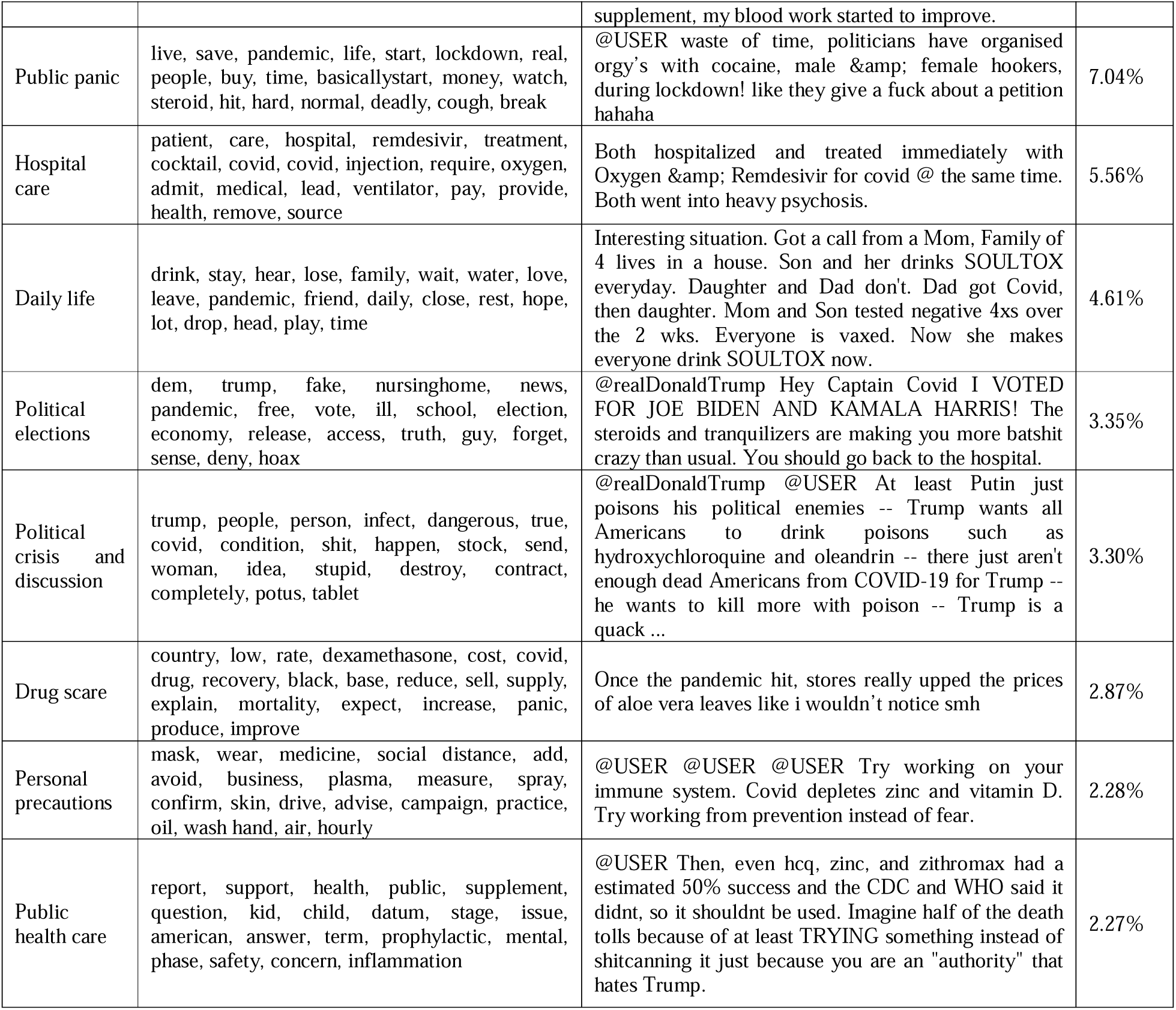
Topic model on drug related Tweets. Username and other sensitive information were masked off using @USER. Public figures such as @realdonaldtrump are shown in their usernames.

| Topic | Keywords | Example | Percent age |
| --- | --- | --- | --- |
| Clinical treatment effect of drugs | treatment, covid, study, drug, effective, trial, hydroxychloroquine, prove, safe, covid, evidence, prevent, clinical, vaccine, recommend, cheap, infection, continue, antiviral, efficacy | @USER Ivermectin is pretty safe but the evidence for it being efficacious against SARS-CoV-2 is lacking. | 13.60% |
| Physical symptoms | day, test, symptom, week, covid, positive, feel, month, steroid, bad, start, med, ago, sick, time, recover, antibiotic, hour, fine, insulin | When I had covid, it was mild fever for a day, & it was gone | 11.84% |
| COVID control | cure, vaccine, spread, control, lie, people, drug, covid, push, approve, hydroxychloroquine, claim, force, medium, ban, talk, science, government, experimental, president | @USER I agree with you re lockdown. Just not on HCQ and vaccines. | 10.36% |
| Causes of death | death, people, die, covid, kill, reason, heart, drug, cancer, cocaine, dead, trust, epidemic, rate, bad, create, attack, sound, result, fentanyl | @USER Of course, if they died 30 minutes after taking fentanyl but had a positive covid test, guess what their official cause of death is listed as? | 9.28% |
| General treatment | doctor, treat, patient, steroid, risk, covid, covid, severe, infection, blood, pill, medication, antibiotic, receive, prescribe, illness, lung, hospitalize, prescription, aspirin | @USER migraines have a very specific causality (i had them for like 20 years), I wonder if the Covid version is one? -- I would try warm compress, NSAIDs, and maybe nasal irrigation with like a neti pot | 8.23% |
| Immune response | virus, system, body, immune, corona, zinc, fight, antibody, cure, immunity, bleach, deficiency, cell, response, kill, covid, inject, injectingdisinfectant, human, boost | In addition, mAbs have been shown to improve survival in patients hospitalized with COVID-19 who have not mounted their own immune response. | 8.14% |
| Daily supplement intake | vitamin, people, level, covid, eat, healthy, cold, flu, catch, protect, food, hand, low, stay, vit, survive, bad, chance, common, worry | @USER regularly take vitamins and a Vitamin D supplement. I started taking the Vitamin D supplement because I wasn't going outside as much at the start of the pandemic. As soon as started taking the Vitamin D supplement, my blood work started to improve. | 7.27% |
| Public panic | live, save, pandemic, life, start, lockdown, real, people, buy, time, basicallystart, money, watch, steroid, hit, hard, normal, deadly, cough, break | @USER waste of time, politicians have organised orgy's with cocaine, male & female hookers, during lockdown! like they give a fuck about a petition hahaha | 7.04% |
| Hospital care | patient, care, hospital, remdesivir, treatment, cocktail, covid, covid, injection, require, oxygen, admit, medical, lead, ventilator, pay, provide, health, remove, source | Both hospitalized and treated immediately with Oxygen & Remdesivir for covid @ the same time. Both went into heavy psychosis. | 5.56% |
| Daily life | drink, stay, hear, lose, family, wait, water, love, leave, pandemic, friend, daily, close, rest, hope, lot, drop, head, play, time | Interesting situation. Got a call from a Mom, Family of 4 lives in a house. Son and her drinks SOULTOX everyday. Daughter and Dad don't. Dad got Covid, then daughter. Mom and Son tested negative 4xs over the 2 wks. Everyone is vaxed. Now she makes everyone drink SOULTOX now. | 4.61% |
| Political elections | dem, trump, fake, nursinghome, news, pandemic, free, vote, ill, school, election, economy, release, access, truth, guy, forget, sense, deny, hoax | @realDonaldTrump Hey Captain Covid I VOTED FOR JOE BIDEN AND KAMALA HARRIS! The steroids and tranquilizers are making you more batshit crazy than usual. You should go back to the hospital. | 3.35% |
| Political crisis and discussion | trump, people, person, infect, dangerous, true, covid, condition, shit, happen, stock, send, woman, idea, stupid, destroy, contract, completely, potus, tablet | @realDonaldTrump @USER At least Putin just poisons his political enemies -- Trump wants all Americans to drink poisons such as hydroxychloroquine and oleandrin -- there just aren't enough dead Americans from COVID-19 for Trump -- he wants to kill more with poison -- Trump is a quack ... | 3.30% |
| Drug scare | country, low, rate, dexamethasone, cost, covid, drug, recovery, black, base, reduce, sell, supply, explain, mortality, expect, increase, panic, produce, improve | Once the pandemic hit, stores really upped the prices of aloe vera leaves like i wouldn't notice smh | 2.87% |
| Personal precautions | mask, wear, medicine, social distance, add, avoid, business, plasma, measure, spray, confirm, skin, drive, advise, campaign, practice, oil, wash hand, air, hourly | @USER @USER @USER Try working on your immune system. Covid depletes zinc and vitamin D. Try working from prevention instead of fear. | 2.28% |
| Public health care | report, support, health, public, supplement, question, kid, child, datum, stage, issue, american, answer, term, prophylactic, mental, phase, safety, concern, inflammation | @USER Then, even hcq, zinc, and zithromax had a estimated 50% success and the CDC and WHO said it didnt, so it shouldnt be used. Imagine half of the death tolls because of at least TRYING something instead of shitcanning it just because you are an "authority" that hates Trump. | 2.27% |

In addition to the overall topic summary, we explored the distribution of the 15 topics for the five drugs. Fig. 4 shows a visualization of the distribution. For ivermectin, the prominent theme was "immune response". In contrast, discussions of remdesivir centered on "hospital care". Hydroxychloroquine received relatively even attention among the three topics "causes of death", "drug scare", and “COVID control”. Vitamin D was frequently mentioned in tweets about "daily life", and the main topics about zinc focused on "hospital care" and "COVID control".

**Fig. 4.**
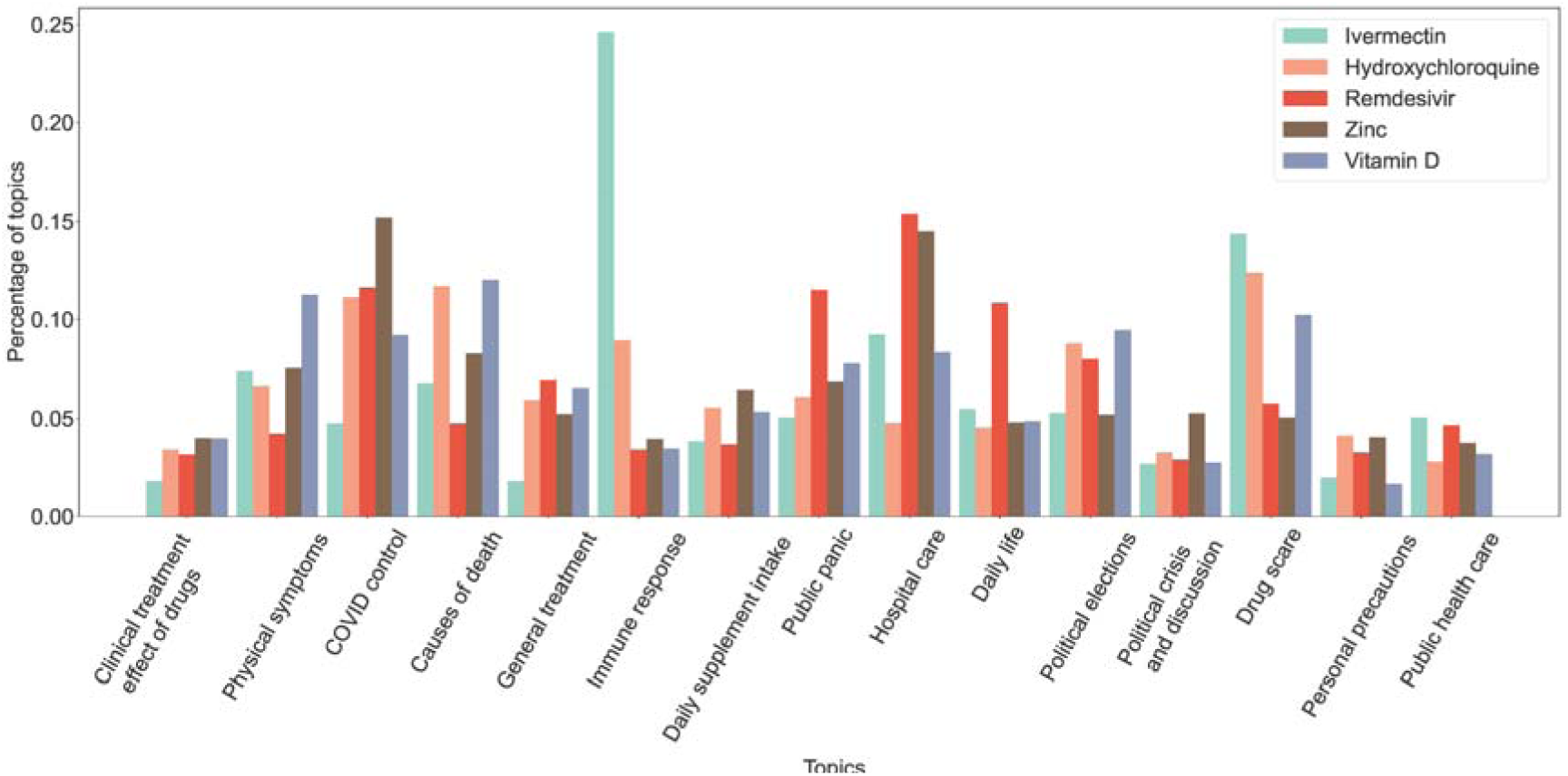
Topic distribution of five top-discussed drugs.

### 3.4 Co-occurrence networks

We visualized the co-occurrence network for drug-drug and drug-symptom relations in Fig. 5. The nodes represent drugs (Table A.3) or symptoms (Table A.4). Node sizes represent node degrees (i.e., the number of linked entities). Edge weights denote the cosine similarity score of two linked nodes.

**Fig. 5.**
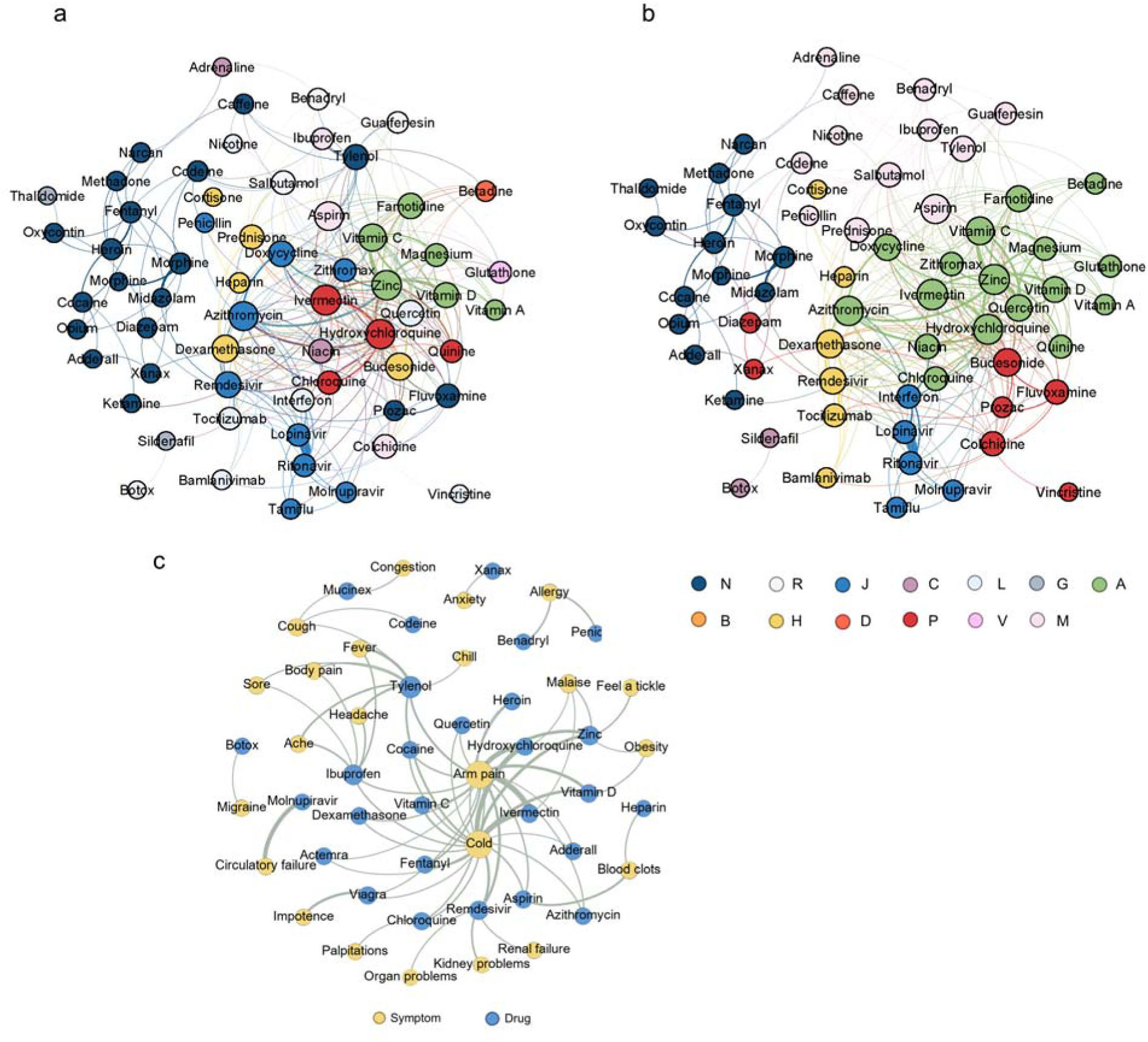
Visualization of drug-related co-occurrence networks by Gephi, including (a) drug-drug associations based on Gephi clustering (τ=0.005), (b) drug-drug associations based on ATC (τ=0.005) and (c) drug-symptom associations (τ=0.05). The color dots on the lower right of the figure represent the ATC categories for Fig. 5b.

#### 3.4.1 Drug-drug network

The origin drug-drug network contained 67 drugs (nodes) with more than 1,000 mentions and 1103 relations (edges) among them. A pre-defined similarity threshold (τ) was established to only visualize relationships with substantial co-occurrence, as measured by cosine similarities exceeding τ. After filtering it with a τ of 0.005, 62 drugs and 317 relations remained in the network. By using Fast Unfolding (Louvian) algorithm,[36] the drugs were clustered in five categories and were colored in Fig. 5a. The same network with drugs colored by ATC classification (12 types) was shown in Fig. 5b for comparison. Drugs in the same group are denoted with the same color. Both figures share similar clustering characters, especially in psychotropic drugs ATC N (e.g., fentanyl, opium, morphine, etc.) and J (e.g., Lopinavir, Ritonavir, Azithromycin). However, drugs in ATC P group (i.e., ivermectin, hydroxychloroquine, quinine and chloroquine) are clustered with the ATC A group in Fig. 5a. The reason may partially lie in the fact that most parasites are intestinal,[37] so most people who need to take anti-parasitic drugs (i.e., ATC P drugs) often present concomitant digestive manifestations,[38] thus necessitating the use of digestive medications (i.e., ATC A drugs), therefore the two drug groups are closely related. Association between some of the significant drug-drug pairs like two Human Immunodeficiency Virus (HIV) protease inhibitors Ritonavir and Lopinavir has been widely studied.[39] Additionally, through the co-occurrence network, we observed several unusual drug pairings, such as midazolam and morphine, salbutamol and prednisone, and zinc and quinine. These strong co-occurrences suggest potential unexplored synergistic effects, adverse reactions, or other public health concerns that warrant further investigation. For instance, we noted a distinct correlation between morphine and midazolam, drugs not typically combined in direct COVID-19 treatment. An analysis of all 376 tweets mentioning both drugs revealed that most discussions focused on end-of-life management for COVID-19 patients and on conspiracy theories about the intentional misuse of these drugs, leading to deaths attributed to causes other than COVID-19.

#### 3.4.2 Drug-symptom network

The drug-symptom network had 136 nodes and 3099 edges and was shown in Fig. 5c. After filtering by τ of 0.05, 50 nodes and 71 edges remained. We observed that the edges often represented symptoms and corresponding treatments, such as Tylenol for fever medication, suggesting the reliability of our association network. We also observed some side effect relations, such as Remdesivir to acute kidney failure[33] and some novel associations receiving no clinical investigation like Molnupiravir to circulatory failure, cocaine to chest cold and vitamin D to malarise. We visualized the top ten closest drugs and symptoms with co-occurrence relationships to the five drugs under investigation (Fig. A.4). These networks revealed the great relevance between hydroxychloroquine, ivermectin and azithromycin from each other. Moreover, Remdesivir was also significantly associated with dexamethasone and Tocilzumab.

## 4. Discussion

This study utilized social media text to develop an NLP-based drug informatic analysis pipeline for assessing public perception of COVID-19-related drugs across time. Leveraging new advances in NLP, we constructed a pretrained language model driven drug entity recognition model and a new targeted sentiment analysis model for polarity prediction of target drugs. This pipeline also includes time trend analysis, topic modeling, and network analysis to explore drug discussions during the pandemic from multiple perspectives. Based on over two years of relevant data, our comprehensive NLP pipeline demonstrates advanced accuracy and completeness in collecting and analyzing data for social media-based drug studies. We will open-source our pipeline, and it can serve as a comprehensive tool to enhance drug safety control and support public health decision-making after the outbreak of infectious diseases.

Compared to traditional pharmaceutical informatics researches, study of drug-related information on social media exhibits distinctive characteristics and advantages. Social media platforms offer real-time and immediate data, enabling the rapid reflection of drug usage patterns and patient feedback, facilitating the prompt identification of potential risks and benefits.[40–42] Furthermore, social media captures viewpoints and experiences of patients, thus furnishing critical insights for the formulation of patient-centered care.[43, 44] For example, understanding patient’s preference on drugs and disease burden can improve the drug development strategies, enabling pharmaceutical companies to better focus on specific drugs that meet patient needs and preferences.[45] In contrast to previous COVID-19 social media studies,[19, 46–48] this work extracted more rigorous data covering a more extended study period and identified five most discussed drugs to be investigated through a fully data-driven method. The substantial volume of social media data allows for large-scale real-time dynamic analysis, and it also covers a broader population than Electronic Health Records (EHRs), which are confined to hospitalized individuals and have restricted access.[49] Social media datasets could also provide large-scale samples for the detection of rare events and the examination of specific population responses, which are challenge in EHR-based analysis.

Sentiment analysis on drugs can highlight patient misconceptions and disagreements about a specific medication, enabling pharmaceutical companies and public health agencies to address public anxiety and reduce misinformation about drugs. Our results confirmed findings from Hua et al.[19] that the public concern and polarity for ivermectin and hydroxychloroquine, which received most social attention, are highly correlated with emotional and political factors, such as personal political orientation, presidential elections, and conspiracy theories. For instance, there was a surge of approximately 200% in acquisitions of medication alternatives such as Hydroxychloroquine within two days subsequent to the press briefing conducted by Donald Trump on March 19, 2020.[1] The topic distribution indicated possible effect or side effect of ivermectin on the immune system and the wide in-hospital treatment use of Remdesivir, but the sentiment analysis showed most opposing stances toward Remdesivir which climbed significantly as the crisis unfolded. It was due to shortages, emergency needs, inefficiency, [50] and potential side effects of Remdesivir like bradycardia,[51] increased risk of hepatic, renal and cardiovascular reactions. [52, 53] Some people even hyped up on Twitter that Remdesivir is approved solely for the purposes of reaping big profits for Anthony Fauci and the democidal cabal that he fronts, bilking the tax payer of billions, and all while quietly euthanizing an unwitting public. Moreover, we also found that daily supplements like zinc and vitamin D did not attract much public attention, but their immune-enhancing properties make them significantly more commended by public than the other three drugs, especially Remdesivir.

It holds immense significance for policy makers and public health agencies to timely track and monitor drug-related concerns on social media, especially for new drug candidates during pandemic. Social media swiftly captures drug-related outbreaks and trends, facilitating rapid policy responses for public safety. It offers direct public interaction, enhancing policymaker understanding of public needs and concerns. This aids in shaping policies aligning with public sentiment, boosting policy acceptability and effectiveness.^40^ Additionally, social media data analysis helps identify drug abuse, adverse reactions, and epidemics, improving health policy planning and resource allocation for evolving health challenges.[54] Our work found that Twitter discussion topics of drugs during COVID-19 were consistent with relevant studies focusing on non-drug COVID-19-related topics.[55–57] Similar to them, this study uncovered public concerns about "public health measures" and "treatment and recovery". In addition, by focusing on drugs, we discovered new drug-specific concerns, such as "drug panic" and "immune response". The focus on "drug panic" may reflect societal uncertainty and anxiety about the drug use during epidemic. Understanding these anxieties can be instrumental in enabling mental health professionals and policymakers to take measures to support mental health and implementing interventions to alleviate anxiety. Cares about the "immune response" may be indicative of public concerns about the immune system, including vaccines and immunotherapies. This can help health agencies better communicate information about vaccinations and immunization support to increase public awareness of immunization.

Many prior studies aimed to detect potential DDI and ADR from social media[58–60] or online literature[61] but largely depended on external vocabulary for keyword-matching and little visualization was performed. This study utilized advanced pretrained language models to identify drug mentions and classify the corresponding sentiments from social media text, ensures the accuracy of information extraction and sentiment prediction. As the pretrained language model is the main NLP structure in our pipeline, it can be easily extended by integrating better large language models (LLMs)[62–64] that have a similar deep learning network structure but with larger parameters, given enough computational resources. Visualization module could illustrate associations between drugs, drug-symptoms pairs, and possible clusters or patterns intuitively and clearly, making it’s easier for researchers to understand and interpret the findings for DDI and ADR.[65] In addition, our co-occurrence network analysis found many widely studied drug-drug and drug-symptom pairs which could verify the reliability of network analysis. The clustering results are consistent with the classification of general clinic (i.e., ATC) to a certain extent, suggesting its potential in capturing similarities and associations between drugs. Notably, we also found many drug pairs with not widely examined associations, such as zinc and quercetin. Their complex (Q/Zn) is considered a potential new drug therapy for improving glycemic control and pulmonary dysfunction in diabetes mellitus,[66] which needs to be further investigated. We found new drug-related associations, such as rheumatoid drugs (hydroxychloroquine, dexamethasone, etc.) may affect COVID-19 treatment due to drug repositioning. Furthermore, networks of top five drugs revealed the significant associations between them such as the co-medication of ivermectin, hydroxychloroquine and azithromycin for COVID-19. Our network analysis also indicated the combination of Remdesivir and Tocilizumab or dexamethasone, and randomized controlled trial showed the efficacy of them for the treatment of severe COVID-19.[67–69]

In essence, the utilization of NLP techniques and network analysis to analyze vast amounts of social media data is an emerging research approach in pharmacovigilance. It holds immense potential in various areas such as the monitoring of ADR, the analysis of drug usage trends, the prediction of epidemics, and the evaluation of drug treatment effects. This novel method has the capacity to serve for pharmaceutical firms, regulatory agencies, and the healthcare fields with more precise and timely information to enhance their efforts in safeguarding public health.

### 4.1 Limitations

Certain limitations apply to this study. First, social media users can’t represent the general population. For example, Twitter users in the U.S. are younger and more likely to be Democrats, and the most prolific 10% of users create 80% of tweets,[70] which may result in bias of our observations. Secondly, although we tried to automate the information extraction with deep learning, we still relied on an empirical lexicon to cluster different concept representations. This allowed us to effectively reduce false positives but not to avoid false negatives. Thirdly, manual checks for symptom recognition suggested that approximately 2-3% of the tweets may still be false positive (e.g., lexical ambiguity like American fever dream), which would lead fake associations, despite the combination of rigorous rules and advanced NLP models based on deep learning. Data accuracy, as well as the reliability of the analysis, are also limited by the authenticity of social media data and the influence of noisy information.

## 5. Conclusion

Our study proposed a pipeline of using social media data and NLP techniques to mine potential drug information, timely track drug-related hot events, facilitate public health stakeholders to conduct reasonable policy enactment, monitor drug public opinion and avoid malignant events in the event of a public health emergency. In addition, it can supplement the existing ADR and DDI databases by constructing multiple medical entity co-occurrence networks to provide real-world clues for future research. Our framework applies not only to COVID-19 but also to other periods of epidemics or major social events. It can also target other public health care foci such as vaccination.

## 6. Summary Table

1. What was already known about the topic?

- The prevalence of the COVID-19 pandemic has induced an urgent demand for efficacious pharmacotherapies.
- Traditional pharmacovigilance mechanisms are plagued by inefficiencies, reporting biases, and a lack of timeliness, thereby lacking comprehensive coverage of the population’s sentiments and experiences.

2. What has this study added to our knowledge?

- The utilization of NLP techniques and network analysis to analyze vast amounts of social media data is an emerging research approach in pharmacovigilance.
- Leveraging new advances in NLP, we could address limitations in the former researches, such as high false positives in information retrieval.
- NLP-based pipeline can be a robust tool for large-scale public health monitoring and can offer valuable supplementary data for traditional epidemiological studies concerning DDI and ADR.

## Data Availability

Data, source code, and pipeline tutorial of this paper are available at https://github.com/zju-liwanxin/covid-twitter-drug.

## Acknowledgements

This research did not receive any specific grant from funding agencies in the public, commercial, or not-for-profit sectors.

## Appendix A. Supplementary data

Supplemental information was submitted in a separate file.

## Author contributions

J.Y., W.L. and X.X. designed the study. W.L and J.Y. drafted the manuscript. Y.H. collected the data, helped draft and revise the manuscript. W.L. performed data and statistical analysis. P.Z. built the NER and TSA models. L.Z. provided critical reviews. All authors reviewed the manuscript. W.L. takes responsibility for the integrity of the work.

## Reference

[1] K. Niburski, O. Niburski. Impact of Trump’s Promotion of Unproven COVID-19 Treatments and Subsequent Internet Trends: Observational Study. J Med Internet Res 22, e20044, doi:10.2196/20044 (2020).

[2] FDA Adverse Event Reporting System (FAERS). <http://www.fda.gov/Drugs/GuidanceComplianceRegulatoryInformation/Surveillance/AdverseDrugEffects/>

[3] R.B. Correia, L. Li, L.M. Rocha, MONITORING POTENTIAL DRUG INTERACTIONS AND REACTIONS VIA 492–503.

[4] R. Harpaz, W. DuMouchel, N.H. Shah, D. Madigan, P. Ryan, C. Friedman. Novel data-mining methodologies for adverse drug event discovery and analysis. Clin. Pharmacol. Ther. 91, 1010–1021, doi:10.1038/clpt.2012.50 (2012).

[5] W.P. Stephenson, M. Hauben. Data mining for signals in spontaneous reporting databases: proceed with caution. Pharmacoepidemiol. Drug Saf. 16, 359–365, doi:10.1002/pds.1323 (2007).

[6] A. Bate, K. Hornbuckle, J. Juhaeri, S.P. Motsko, R.F. Reynolds. Hypothesis-free signal detection in healthcare databases: finding its value for pharmacovigilance. Therapeutic advances in drug safety 10, 2042098619864744, doi:10.1177/2042098619864744 (2019).

[7] A. Bate, S.J. Evans. Quantitative signal detection using spontaneous ADR reporting. Pharmacoepidemiol. Drug Saf. 18, 427–436, doi:10.1002/pds.1742 (2009).

[8] M.D. Rawlins. Spontaneous reporting of adverse drug reactions. I: the data. Br J Clin Pharmacol 26, 1–5, doi:10.1111/j.1365-2125.1988.tb03356.x (1988).

[9] A. Nikfarjam, A. Sarker, K. O’Connor, R. Ginn, G. Gonzalez. Pharmacovigilance from social media: mining adverse drug reaction mentions using sequence labeling with word embedding cluster features. J Am Med Inform Assoc 22, 671–681, doi:10.1093/jamia/ocu041 (2015).

[10] Z. Yin, L.M. Sulieman, B.A. Malin. A systematic literature review of machine learning in online personal health data. J Am Med Inform Assoc 26, 561–576, doi:10.1093/jamia/ocz009 (2019).

[11] S. Rees, S. Mian, N. Grabowski. Using social media in safety signal management: is it reliable? Therapeutic advances in drug safety 9, 591–599, doi:10.1177/2042098618789596 (2018).

[12] N. Mekawie, A. Hany. Understanding the factors driving consumers’ purchase intention of over the counter medications using social media advertising in Egypt:(A Facebook advertising application for cold and Flu products). Procedia Computer Science 164, 698–705 (2019).

[13] M. Alshareef, A. Alotiby. Prevalence and Perception Among Saudi Arabian Population About Resharing of Information on Social Media Regarding Natural Remedies as Protective Measures Against COVID-19. International journal of general medicine 14, 5127–5137, doi:10.2147/IJGM.S326767 (2021).

[14] A.J. Lazard. Social media message designs to educate adolescents about e-cigarettes. J. Adolesc. Health 68, 130–137 (2021).

[15] J. Wu, X. Wu, Y. Hua, S. Lin, Y. Zheng, J. Yang, Exploring Social Media for Early Detection of Depression in COVID-19 Patients, Proceedings of the ACM Web Conference 2023, Association for Computing Machinery, 2023, pp. 3968–3977.

[16] M.T. Osborne, E. Kenah, K. Lancaster, J. Tien. Catch the tweet to fight the flu: Using Twitter to promote flu shots on a college campus. Journal of American college health : J of ACH 71, 2470–2484, doi:10.1080/07448481.2021.1973480 (2023).

[17] H. Jiang, Y. Hua, D. Beeferman, D. Roy. Annotating the Tweebank Corpus on Named Entity Recognition and Building NLP Models for Social Media Analysis. arXiv preprint arXiv:.07281, doi: https://arxiv.org/abs/2201.07281 (2022).

[18] P. Zhou, Z. Wang, D. Chong, Z. Guo, Y. Hua, Z. Su, Z. Teng, J. Wu, J. Yang. *METS-CoV: A Dataset of Medical Entity and Targeted Sentiment on COVID-19 Related Tweets* in *Advances in Neural Information Processing Systems (NeurIPS)*.

[19] Y. Hua, H. Jiang, S. Lin, J. Yang, J.M. Plasek, D.W. Bates, L. Zhou. Using Twitter data to understand public perceptions of approved versus off-label use for COVID-19-related medications. J Am Med Inform Assoc 29, 1668–1678, doi:10.1093/jamia/ocac114 (2022).

[20] J. Wu, L. Wang, Y. Hua, M. Li, L. Zhou, D.W. Bates, J. Yang. Trend and Co-occurrence Network of COVID-19 (2023).

[21] E. Chen, K. Lerman, E. Ferrara. Tracking Social Media Discourse About the COVID-19 Pandemic: Development of a Public Coronavirus Twitter Data Set. JMIR Public Health Surveill 6, e19273, doi:10.2196/19273 (2020).

[22] J. Devlin, M.-W. Chang, K. Lee, K. Toutanova. Bert: Pre-training of deep bidirectional transformers for language understanding. arXiv preprint arXiv:1810.04805 (2018).

[23] R. Rehurek, P. Sojka. *Software framework for topic modelling with large corpora* in *In Proceedings of the LREC 2010 workshop on new challenges for NLP frameworks*. (Citeseer).

[24] Y. Guo, Y. Zhang, T. Lyu, M. Prosperi, F. Wang, H. Xu, J. Bian. The application of artificial intelligence and data integration in COVID-19 studies: a scoping review. Journal of the American Medical Informatics Association 28, 2050–2067, doi:10.1093/jamia/ocab098 (2021).

[25] K. Stevens, P. Kegelmeyer, D. Andrzejewski, D. Buttler. *Exploring topic coherence over many models and many topics* in *Proceedings of the 2012 joint conference on empirical methods in natural language processing and computational natural language learning*. 952–961.

[26] Gephi, <https://gephi.org/about/> (2017).

[27] the Anatomical Therapeutic Chemical Classification System (ATC) from WHO, <https://www.whocc.no/atc_ddd_index/.>

[28] Y. Hua, S. Lin, M. Li, Y. Zhang, P. Zhou, Y.-C. Lo, L. Zhou, J. Yang. Streamlining Social Media Information Retrieval for Public Health Research with Deep Learning. Journal of the American Medical Informatics Association ocae118 (2024).

[29] F. Rahutomo, T. Kitasuka, M. Aritsugi. *Semantic cosine similarity* in *The 7th international student conference on advanced science and technology ICAST*. 1.

[30] WHO. *Coronavirus disease* (*COVID-19*) *Situation dashboard*, *as of January*, *7 2021 [EB/OL].* (2022-01-07), <https//covid19.who.int.>

[31] Twitter: Most Users by Country [EB/OL]. <https://www.statista.com/statistics/242606/number-of-active-twitter-users-in-selected-countries/> (2021).

[32] K. Ansems, F. Grundeis, K. Dahms, A. Mikolajewska, V. Thieme, V. Piechotta, M.I. Metzendorf, M. Stegemann, C. Benstoem, F. Fichtner. Remdesivir for the treatment of COVID-19. Cochrane Database Syst Rev 8, CD014962, doi:10.1002/14651858.CD014962 (2021).

[33] M.M. Rahimi, E. Jahantabi, B. Lotfi, M. Forouzesh, R. Valizadeh, S. Farshid. Renal and liver injury following the treatment of COVID-19 by remdesivir. Journal of Nephropathology 10, 1–4 (2021).

[34] M.A.R. Guerra, C. Mendoza, S. Kandhi, H. Sun, M. Saad, T. Vittorio. Cardiac arrhythmia related to remdesivir in COVID-19. ISMMS Journal of Science and Medicine 1 (2021).

[35] E. Sneij, V. Kohli, S.A. Al-Adwan, A. Mealor. Remdesivir Causing Profound Bradycardia. Journal of the American College of Cardiology 77, 2037–2037, doi:10.1016/s0735-1097(21)03393-3 (2021).

[36] V.D. Blondel, J.-L. Guillaume, R. Lambiotte, E. Lefebvre. Fast unfolding of communities in large networks. Journal of Statistical Mechanics: Theory and Experiment 2008, P10008 (2008).

[37] A. Naveed, S. Abdullah. Impact of parasitic infection on human gut ecology and immune regulations. Translational Medicine Communications 6, 11, doi:10.1186/s41231-021-00091-4 (2021).

[38] I.C.M. Sey, A.M. Ehimiyein, C. Bottomley, E.M. Riley, J.P. Mooney. Does Malaria Cause Diarrhoea? A Systematic Review. Frontiers in medicine 7, 589379, doi:10.3389/fmed.2020.589379 (2020).

[39] R.S. Cvetkovic, K.L. Goa. Lopinavir/ritonavir: a review of its use in the management of HIV infection. Drugs 63,

[40] G. Eysenbach. Infodemiology and infoveillance: framework for an emerging set of public health informatics methods to analyze search, communication and publication behavior on the Internet. J Med Internet Res 11, e11, doi:10.2196/jmir.1157 (2009).

[41] J. Chen, Y. Wang. Social Media Use for Health Purposes: Systematic Review. J Med Internet Res 23, e17917, doi:10.2196/17917 (2021).

[42] A. Gupta, R. Katarya. Social media based surveillance systems for healthcare using machine learning: A systematic review. J. Biomed. Inform. 108, 103500, doi:10.1016/j.jbi.2020.103500 (2020).

[43] L. McDonald, B. Malcolm, S. Ramagopalan, H. Syrad. Real-world data and the patient perspective: the PROmise of social media? BMC Med. 17, 11, doi:10.1186/s12916-018-1247-8 (2019).

[44] S.R. Kudchadkar, C.L. Carroll. Using Social Media for Rapid Information Dissemination in a Pandemic: #PedsICU and Coronavirus Disease 2019. Pediatr. Crit. Care Med. 21, e538–e546, doi:10.1097/PCC.0000000000002474 (2020).

[45] A.L. Schmidt, R. Rodriguez-Esteban, J. Gottowik, M. Leddin. Applications of quantitative social media listening to patient-centric drug development. Drug discovery today 27, 1523–1530, doi:10.1016/j.drudis.2022.01.015 (2022).

[46] M. Li, Y. Hua, Y. Liao, L. Zhou, X. Li, L. Wang, J. Yang. Tracking the Impact of COVID-19 and Lockdown Policies on Public Mental Health Using Social Media: Infoveillance Study. J Med Internet Res 24, e39676, doi:10.2196/39676 (2022).

[47] J.C. Lyu, E.L. Han, G.K. Luli. COVID-19 Vaccine-Related Discussion on Twitter: Topic Modeling and Sentiment Analysis. J Med Internet Res 23, e24435, doi:10.2196/24435 (2021).

[48] M. Al-Ramahi, A. Elnoshokaty, O. El-Gayar, T. Nasralah, A. Wahbeh. Public Discourse Against Masks in the COVID-19 Era: Infodemiology Study of Twitter Data. JMIR Public Health Surveill 7, e26780, doi:10.2196/26780 (2021).

[49] J. Wu, X. Liu, M. Li, W. Li, Z. Su, S. Lin, L. Garay, Z. Zhang, Y. Zhang, Q. Zeng, J. Shen, C. Yuan, J. Yang. Clinical Text Datasets for Medical Artificial Intelligence and Large Language Models — A Systematic Review. NEJM AI 1, AIra2400012, doi:doi:10.1056/AIra2400012 (2024).

[50] K. Hagman, M. Hedenstierna, J. Widaeus, E. Arvidsson, B. Hammas, L. Grillner, J. Jakobsson, P. Gille-Johnson, J. Ursing. Effects of remdesivir on SARS-CoV-2 viral dynamics and mortality in viraemic patients hospitalized for COVID-19. J. Antimicrob. Chemother. 78, 2735–2742, doi:10.1093/jac/dkad295 (2023).

[51] Y. Ishisaka, T. Aikawa, A. Malik, P.N. Kampaktsis, A. Briasoulis, T. Kuno. Association of Remdesivir use with bradycardia: A systematic review and meta-analysis. J. Med. Virol. 95, e29018, doi:10.1002/jmv.29018 (2023).

[52] H.A. Blair. Remdesivir: A Review in COVID-19. Drugs 83, 1215–1237, doi:10.1007/s40265-023-01926-0 (2023).

[53] T. Akhvlediani, R. Bernard-Valnet, S.P. Dias, R. Eikeland, B. Pfausler, J. Sellner. Neurological side effects and drug interactions of antiviral compounds against SARS-CoV-2. Eur. J. Neurol. 30, 3904–3912, doi:10.1111/ene.16017 (2023).

[54] J. van Stekelenborg, J. Ellenius, S. Maskell, T. Bergvall, O. Caster, N. Dasgupta, J. Dietrich, S. Gama, D. Lewis, V. Newbould, S. Brosch, C.E. Pierce, G. Powell, A. Ptaszynska-Neophytou, A.F.Z. Wisniewski, P. Tregunno, G.N. Noren, M. Pirmohamed. Recommendations for the Use of Social Media in Pharmacovigilance: Lessons from IMI WEB-RADR. Drug Saf 42, 1393–1407, doi:10.1007/s40264-019-00858-7 (2019).

[55] S. Boon-Itt, Y. Skunkan. Public Perception of the COVID-19 Pandemic on Twitter: Sentiment Analysis and Topic Modeling Study. JMIR Public Health Surveill 6, e21978, doi:10.2196/21978 (2020).

[56] R. Chandrasekaran, V. Mehta, T. Valkunde, E. Moustakas. Topics, Trends, and Sentiments of Tweets About the COVID-19 Pandemic: Temporal Infoveillance Study. J Med Internet Res 22, e22624, doi:10.2196/22624 (2020).

[57] A. Boukobza, A. Burgun, B. Roudier, R. Tsopra. Deep Neural Networks for Simultaneously Capturing Public Topics and Sentiments During a Pandemic: Application on a COVID-19 Tweet Data Set. JMIR Med Inform 10, e34306, doi:10.2196/34306 (2022).

[58] H. Yang, C.C. Yang. *Harnessing Social Media for Drug-Drug Interactions Detection* in 2013 *IEEE International*

[59] L. Xia, G.A. Wang, W. Fan. *A deep learning based named entity recognition approach for adverse drug events identification and extraction in health social media* in *International Conference on Smart Health*. 237–248 (Springer).

[60] P.M. Coloma, B. Becker, M.C. Sturkenboom, E.M. van Mulligen, J.A. Kors. Evaluating Social Media Networks in Medicines Safety Surveillance: Two Case Studies. Drug Saf 38, 921–930, doi:10.1007/s40264-015-0333-5 (2015).

[61] Y. Lu, D. Shen, M. Pietsch, C. Nagar, Z. Fadli, H. Huang, Y.C. Tu, F. Cheng. A novel algorithm for analyzing drug-drug interactions from MEDLINE literature. Sci. Rep. 5, 17357, doi:10.1038/srep17357 (2015).

[62] T.B. Brown, B. Mann, N. Ryder, M. Subbiah, J. Kaplan, P. Dhariwal, A. Neelakantan, P. Shyam, G. Sastry, A. Askell, S. Agarwal, A. Herbert-Voss, G. Krueger, T. Henighan, R. Child, A. Ramesh, D.M. Ziegler, J. Wu, C. Winter, C. Hesse, M. Chen, E. Sigler, M. Litwin, S. Gray, B. Chess, J. Clark, C. Berner, S. McCandlish, A. Radford, I. Sutskever, D. Amodei. Language Models are Few-Shot Learners. Adv. Neural Inf. Process. Syst. abs/2005.14165, 1877–1901 (2020).

[63] K. Singhal, S. Azizi, T. Tu, S.S. Mahdavi, J. Wei, H.W. Chung, N. Scales, A. Tanwani, H. Cole-Lewis, S. Pfohl, P. Payne, M. Seneviratne, P. Gamble, C. Kelly, A. Babiker, N. Schärli, A. Chowdhery, P. Mansfield, D. Demner-Fushman, Y.A.B. Agüera, D. Webster, G.S. Corrado, Y. Matias, K. Chou, J. Gottweis, N. Tomasev, Y. Liu, A. Rajkomar, J. Barral, C. Semturs, A. Karthikesalingam, V. Natarajan. Large language models encode clinical knowledge. Nature 620, 172–180, doi:10.1038/s41586-023-06291-2 (2023).

[64] J. Wu, X. Wu, Z. Qiu, M. Li, S. Lin, Y. Zhang, Y. Zheng, C. Yuan, J. Yang. Large language models leverage external knowledge to extend clinical insight beyond language boundaries. Journal of the American Medical Informatics Association, doi:10.1093/jamia/ocae079 (2024).

[65] R. Wang, S. Li, L. Cheng, M.H. Wong, K.S. Leung. Predicting associations among drugs, targets and diseases by tensor decomposition for drug repositioning. BMC Bioinformatics 20, 628, doi:10.1186/s12859-019-3283-6 (2019).

[66] M.S. Refat, R.Z. Hamza, A.M.A. Adam, H.A. Saad, A.A. Gobouri, F.S. Al-Harbi, F.A. Al-Salmi, T. Altalhi, S.M. El-Megharbel. Quercetin/Zinc complex and stem cells: A new drug therapy to ameliorate glycometabolic control and pulmonary dysfunction in diabetes mellitus: Structural characterization and genetic studies. PLoS One 16, e0246265, doi:10.1371/journal.pone.0246265 (2021).

[67] A.T.M. Mohiuddin Chowdhury, A. Kamal, K.U. Abbas, S. Talukder, M.R. Karim, M.A. Ali, M. Nuruzzaman, Y. Li, S. He. Efficacy and Outcome of Remdesivir and Tocilizumab Combination Against Dexamethasone for the Treatment of Severe COVID-19: A Randomized Controlled Trial. Front. Pharmacol. 13, 690726, doi:10.3389/fphar.2022.690726 (2022).

[68] A. Marrone, R. Nevola, A. Sellitto, D. Cozzolino, C. Romano, G. Cuomo, C. Aprea, M.X.P. Schwartzbaum, C. Ricozzi, S. Imbriani, L. Rinaldi, K. Gjeloshi, A. Padula, R. Ranieri, C. Ruosi, L.A. Meo, M. Abitabile, F. Cinone, C. Carusone, L.E. Adinolfi. Remdesivir Plus Dexamethasone Versus Dexamethasone Alone for the Treatment of Coronavirus Disease 2019 (COVID-19) Patients Requiring Supplemental O2 Therapy: A Prospective Controlled Nonrandomized Study. Clin. Infect. Dis. 75, e403–e409, doi:10.1093/cid/ciac014 (2022).

[69] S.B. Gressens, V. Esnault, N. De Castro, P. Sellier, D. Sene, L. Chantelot, B. Hervier, C. Delaugerre, S. Chevret, J.M. Molina, C.g. Saint-Louis. Remdesivir in combination with dexamethasone for patients hospitalized with COVID-19: A retrospective multicenter study. PLoS One 17, e0262564, doi:10.1371/journal.pone.0262564 (2022).

[70] S. Wojcik, A. Hughes. Sizing up Twitter users. PEW research center 24 (2019).

