## Supplementary materials for "Characterizing Public Sentiments and Drug Interactions during COVID-19: A Pretrained Language Model and Network Analysis of Social Media Discourse"

### Table A.1 Comparison of NER + lexicon and pure lexicon-based extraction

| Accuracy | Entity | NER + lexicon | lexicon-based |
| --- | --- | --- | --- |
| Drug | Ivermectin | 100% | 100% |
|  | Hydroxychloroquine | 100% | 100% |
|  | Remdesivir | 100% | 100% |
|  | Zinc | 100% | 98% |
|  | Vitamin D | 100% | 98% |
| Symptom | Fever | 95% | 60% |
|  | Cough | 98% | 92% |
|  | Headache | 97% | 90% |
|  | Allergy | 99% | 65% |
|  | Blood clots | 100% | 85% |
| Average | - | 97.8% | 89% |

We used two methods (i.e., NER + lexicon and lexicon-based) to randomly extract 50 tweets for each top five entities and calculated the recognition accuracy.

### Table A.2 Statistical results of medical entity recognition

| Type | Number of types | Number of entities |
| --- | --- | --- |
| Person | 6,383,455 | 54,829,602 |
| Organization | 1,970,671 | 35,572,099 |
| Location | 968,013 | 41,669,407 |
| Drug | 166,288 | 2,819,706 |
| Disease | 153,685 | 10,415,858 |
| Vaccine-related | 242,468 | 6,841,926 |
| Symptom | 498,494 | 6,176,421 |

### Table A.3 67 top drug entities (frequency >1000)

| Drug | ATC^[1]^ | Lexicon^[2]^ | Freq |
| --- | --- | --- | --- |
| Ivermectin | P | Ivermectin\|Stromectol | 345098 |
| Hydroxychloroquine | P | Hydroxychloroquine\|HCQ\|plaquenil | 267108 |
| Zinc | A | zinc | 91437 |
| Vitamin D | A | vitamin d\|Vitamin D2\|Vitamin D3\|vd3\|vd2\|Calciferols | 87259 |
| Remdesivir | J | Remdesivir\|velkury | 66920 |
| Tylenol | N | tylenol\|acetaminophen\|Paracetamol\|APAP\|Panadol\|Calpol | 41112 |
| Vitamin C | A | Vitamin C\|vit c | 41026 |
| Fentanyl | N | Fentanyl\|Actiq\|Duragesic\|Sublimaze, | 33174 |
| Cocaine | N | Cocaine | 21778 |
| Aspirin | M | aspirin | 18881 |
| Chloroquine | P | Chloroquine | 18456 |
| Ibuprofen | M | ibuprofen\|advil\|motrin\|Nurofen\|brufen | 17925 |
| Heroin | N | Heroin | 14507 |
| Dexamethasone | H | dexamethasone | 14504 |
| Quercetin | L | Quercetin | 12732 |
| Azithromycin | J | azithromycin | 12065 |
| Sildenafil | G | viagra\|Sildenafil | 9254 |
| Nicotine | L | nicotine | 8943 |
| Caffeine | N | Caffeine | 8109 |
| Tocilizumab | L | actemra\|Tocilizumab\|RoActemra | 6879 |
| Botox | L | botox | 6603 |
| Adderall | N | adderall | 6231 |
| Adrenaline | C | Adrenaline\|epipen\|Epinephrine\|Adrenaclick | 5698 |
| Penicillin | J | penicillin | 5328 |
| Molnupiravir | J | Molnupiravir | 5326 |
| Midazolam | N | Midazolam | 5205 |
| Testosterone | G | testosterone | 5088 |
| Magnesium | A | magnesium | 5046 |
| Thalidomide | G | Thalidomide\|Contergan\|Thalomid\|Talidex | 5015 |
| Tamiflu | J | tamiflu\|Oseltamivir | 4788 |
| Prednisone | H | prednisone | 4570 |
| Fluvoxamine | N | Fluvoxamine | 4232 |
| Morphine | N | Morphine | 4013 |
| Niacin | C | niacin\|nicotinic acid | 3739 |
| Budesonide | H | budesonide | 3641 |
| Doxycycline | J | doxycycline | 3566 |
| Quinine | P | Quinine | 3526 |
| Interferon | L | Interferon | 3274 |
| Midazolam | N | midazolam | 3227 |
| Xanax | N | xanax | 2969 |
| Benadryl | R | benadryl\|Diphenhydramine | 2919 |
| Opium | N | Opium | 2837 |
| Oxycontin | N | oxycontin\|Roxicodone | 2741 |
| Morphine | N | morphine | 2664 |
| Famotidine | A | famotidine\|Pepcid | 2643 |
| Guaifenesin | R | mucinex\|Guaifenesin | 2527 |
| Zithromax | J | Zithromax\|azithromycin | 2040 |
| Ketamine | N | Ketamine | 1906 |
| Betadine | D | betadine\|Povidone-iodine | 1891 |
| Salbutamol | R | albuterol\|Salbutamol | 1799 |
| Narcan | N | naloxone\|Narcan | 1721 |
| Heparin | B | Heparin | 1720 |
| Vincristine | L | vincristine\|Oncovin\|Vincasar | 1625 |
| Amphotericin B | J | amphotericin b\| Fungizone | 1576 |
| Prozac | N | prozac\|Fluoxetine\|Animex-On\|Sarafem\|Adofen\|Deprex | 1555 |
| Glutathione | V | Glutathione | 1367 |
| Lopinavir | J | lopinavir\|ritonavir | 1331 |
| Diazepam | N | valium\|Diazepam | 1281 |
| Menthol | N | Menthol | 1280 |
| Vaseline | D | Vaseline | 1245 |
| Bamlanivimab | L | bamlanivimab | 1181 |
| Codeine | N | codeine | 1154 |
| Colchicine | M | colchicine\|Colcrys | 1150 |
| Ritonavir | J | ritonavir | 1136 |
| Vitamin A | A | vitamin a\|Retinol | 1079 |
| Methadone | N | Methadone\|Dolophine | 1043 |
| Cortisone | H | Cortisone | 1038 |

### Table A.4 69 top symptom entities(frequency>250)

| Symptom | Lexicon | Freq |
| --- | --- | --- |
| Fever | fever\|fevers\|feverish\|fevered\|high temperature\|temperature\|mild temperature\|elevated temperature | 19464 |
| Cough | cough\|coughing\|coughed\|coughs\|dry cough\|chronic cough | 15690 |
| Headache | headache\|headaches\|head ache\|head cold\|head colds\|head hurt\|head hurts | 13049 |
| Allergy | allergies\|allergic\|allergy\|allergic reaction\|allergic reactions\|seasonal allergies\|allergens\|drug allergies\|anaphylaxis | 11399 |
| Blood clots | blood clots\|blood clotting\|blood clot\|clots\|clotting\|clot\|micro clots\|blood clots in the lungs\|clotting issues | 11376 |
| Dyspnea | respiratory distress\|respiratory issues\|respiratory problems\|respiratory infection\|acute respiratory syndrome\|upper respiratory infection\|respiratory depression\|respiratory illness\|respiratory symptoms\|respiratory infections\|shortness of breath\|can't breathe\|can ' t breathe\|breathing issues\|breathing problems\|breathlessness\|difficulty breathing\|couldn ' t breathe\|short of breath\|couldn't breathe\|breathing difficulties\|breathless\|struggling to breathe\|trouble breathing\|having trouble breathing\|can ' t breath\|can't breath\|have trouble breathing\|couldn't breath\|couldn ' t breath\|hard to breathe\|breathing problem\|out of breath\|breathing difficulty\|had trouble breathing\|unable to breathe\|cant breathe\|difficulty in breathing\|hard time breathing\|cannot breathe\|gasping for breath\|not being able to breathe\|could not breathe\|struggling to breath\|bad breath\|breathing trouble\|hard to breath\|troubles in breathing\|breathing issue\|problems breathing\|struggle to breathe\|stop breathing\|cant breath\|labored breathing\|dyspnea\|gasping | 11304 |
| Fatigue | fatigue\|fatigued\|tired\|tiredness\|weak\|weakness\|muscle weakness\|no energy\|low energy\|chronic fatigue | 9031 |
| Ache | pain\|pains\|ache | 6459 |
| Pain in throat | bad throat\|dry throat\|itchy throat\|scratchy throat\|soar throat\|sore throat\|sore throats\|sorethroat\|sour throat\|throat hurt\|throat hurting\|throat hurts\|throat infection\|throat infections\|throat irritation\|throat is sore\|throat issues\|throat pain\|throat sore\|throat tickle\|throat was sore\|tickle in my throat\|tickly throat\|viral sore throat | 4659 |
| Rhinorrhea | runny nose\|stuffy nose\|running nose\|blocked nose\|runny noses\|sniffles\|sniffle\|sniffling\|sniffing\|post nasal drip\|nasal drip | 4383 |
| Malaise | sick\|sickness\|malaise\|discomfort | 4028 |
| Body pain | body aches\|body pain\|body ache\|body pains\|bodyache\|body hurts\|muscle aches\|muscle pain\|muscle soreness\|joint pain\|neck pain\|back pain\|backache | 3938 |
| Chest pain | chest infection\|chest pain\|chest pains\|chest congestion\|chest infections\|chest tightness\|chest cold\|tight chest\|chesty cough\|chest hurts | 2883 |
| Cold | chest cold\|cold\|cold sores\|cold sweat\|cold sweats\|\|colds\|head cold\|head colds\|headcold\|hot and cold\|sinus cold | 2745 |
| Chill | chills\|chill\|chillin\|shivering\|shivers | 2733 |
| Obesity | obesity\|fat | 2642 |
| Taste sense altered | loss of taste and smell\|loss of taste\|no taste\|lost taste and smell\|loss of taste smell and apatite\|no taste or smell\|loss of taste / smell\|loss of smell and taste\|can't taste\|can ' t taste\|loss of taste smell\|lost taste\|loss of smell / taste\|lost my taste and smell\|lost my taste\|lost my sense of taste\|lost my sense of taste and smell\|lost smell and taste\|couldn't taste\|lost taste / smell\|lost my sense of smell and taste\|lost taste smell\|couldn ' t taste | 2342 |
| Anxiety | anxiety\|anxiety attack\|anxiety attacks\|anxiety disorder\|anxiety issues\|anxious | 2328 |
| Inflammation | acute inflammation\|brain inflammation\|chronic inflammation\|heart inflammation\|hyper - inflammation\|hyper inflammation\|hyperinflammation\|inflamation\|inflamed\|inflamed lungs\|inflammation\|inflammation in his lungs\|inflammation in my lungs\|inflammation in the heart\|inflammation in the lungs\|inflammation of the lungs\|joint inflammation\|liver inflammation\|lung inflammation\|lungs are inflamed\|pulmonary inflammation\|respiratory inflammation\|systemic inflammation\|vascular inflammation | 2215 |
| Kidney problems | acute kidney failure\|acute kidney injury\|kidney and liver damage\|kidney and liver failure\|kidney damage\|kidney failure\|kidney failures\|kidney infection\|kidney injuries\|kidney injury\|kidney issues\|kidney liver failure\|kidney pain\|kidney problems\|kidney stones\|kidneys are failing\|kidneys fail\|kidneys failed\|kidneys shut down\|kidneys to fail\|kidneys to shut down\|liver and kidney damage\|liver and kidney failure\|liver and kidney problems\|shuts down kidneys | 2176 |
| Sore | arm is a little sore\|arm is sore\|arm is still sore\|arm sore\|arm soreness\|arm was sore\|body soreness\|cold sores\|injection site soreness\|mouth sores\|muscle soreness\|sore\|sore arm\|sore arms\|sore back\|sore body\|sore chest\|sore eyes\|sore head\|sore joints\|sore muscles\|sore shoulder\|soreness\|soreness in my arm\|sores | 2124 |
| Insomnia | insomnia\|insomniac\|bad sleep\|can ' t sleep\|cannot sleep\|cant sleep\|can't sleep\|couldn ' t sleep\|couldn't sleep\|had trouble sleeping\|hard to sleep\|have trouble sleeping\|having trouble sleeping\|lack of sleep\|no sleep\|not sleeping\|poor sleep\|sleep deprivation\|sleep issues\|sleep problems\|sleepless\|sleeplessness\|trouble sleeping\|unable to sleep\|sleep issues\|sleep problems | 2108 |
| Sneezing | sneezing\|sneeze\|sneezed\|sneezes\|sneezy | 2061 |
| Lung problems | lung damage\|lung infection\|lung inflammation\|lung issues\|lung problems\|lung injury\|lung injuries\|damaged lungs\|lungs infection\|inflammation in the lungs\|lung involvement\|lungs fill with fluid\|fluid in the lungs | 2019 |
| Heart problems | enlarged heart\|enlarged hearts\|heart and lung issues\|heart complications\|heart inflammation\|heart injury\|heart issue\|heart issues\|heart pain\|heart problem\|heart problems\|heart probs\|heart trouble\|heartache\|heartworms\|inflammation in the heart\|heart burn\|heartburn | 1950 |
| Nausea | nausea\|nauseated\|nauseous | 1926 |
| Arm pain | sore arm\|covid arm\|arm pain\|arm soreness\|arm hurts\|arm hurt\|broken arm\|arm was sore\|arm is sore\| | 1922 |
| Congestion | chest congestion\|congested\|congested chest\|congested nose\|congestion\|head congestion\|lung congestion\|nasal congestion\|sinus congestion | 1797 |
| Sinus infection | sinus infection | 1679 |
| Diarrhea | diarrhoea\|diarrhea | 1638 |
| Hypoxia | lack of oxygen\|low oxygen\|shortage of oxygen\|low oxygen levels\|oxygen deprivation\|hypoxia\|hypoxic | 1625 |
| Concentration problems | brain - fog\|brain fog\|brainfog\|foggy brain | 1624 |
| Dipression | depressed\|depression\|respiratory depression\|seasonal depression | 1584 |
| Anosmia | loss of smell\|can't smell\|lost my sense of smell\|can ' t smell\|no smell\|sense of smell\|lost smell\|couldn't smell\|lost sense of smell\|couldn ' t smell\|lost my smell\|anosmia | 1527 |
| Palpitations | abnormal heart rhythm\|abnormal heart rhythms\|elevated heart rate\|heart arrhythmia\|heart arrhythmias\|heart arrythmia\|heart irregularities\|heart race\|heart racing\|heart rate spikes\|heart rhythm issues\|heart rhythm problems\|high heart rate\|irregular heart beat\|irregular heart rate\|irregular heart rates\|irregular heart rhythm\|irregular heart rhythms\|irregular heartbeat\|irregular heartbeats\|racing heart\|rapid heart rate\|rapid heartbeat\|heart palpitations | 1439 |
| Chronic pain | chronic back pain\|chronic intractable pain\|chronic pain | 1433 |
| Vomiting | vomiting\|vomit\|vomited\|throwing up\|throw up | 1351 |
| Organ problems | damaged organs\|multi - organ failure\|multi organ failure\|multi symptom organ shutdown\|multiorgan failure\|multiple organ failure\|organ damage\|organ failure\|organ failures\|organ impairment\|organs shut down\|organs shutting down | 1261 |
| Dizziness | dizzy\|dizziness\|vertigo\|lightheaded | 1246 |
| Exhausted | exhausted\|exhausting\|exhaustion\|heat exhaustion | 1191 |
| Liver problems | elevated liver enzymes\|fatty liver\|kidney and liver damage\|kidney and liver failure\|kidney liver failure\|liver and kidney damage\|liver and kidney failure\|liver and kidney problems\|liver damage\|liver failure\|liver inflammation\|liver injury\|liver issues\|liver problem\|liver problems\|liver toxicity | 1131 |
| Cytokine storm | cytokine storm\|cytokine storms\|cytokines storm | 1058 |
| Ear problems | ear infection\|hearing loss\|ear infections\|earache\|ear ache\|tinnitus | 1048 |
| Seizures | seizures\|muscle spasms\|agonising cramps\|cramping\|cramps\|leg cramps\|menstrual cramps\|muscle cramps\|period cramps\|stomach cramps | 1000 |
| Dehydration | dehydration\|dehydrated | 984 |
| Rash | covid rash\|itchy rash\|rash\|rashes\|skin rash\|skin rashes | 948 |
| Migraine | chronic migraines\|migraine\|migraines | 893 |
| Brain problems | brain bleeds\|brain damage\|brain damaged\|brain dead\|brain hurt\|brain hurts\|brain inflammation\|brain injuries\|brain injury\|brain issues\|brain shrinkage\|brain swelling\|brain zaps\|brain hemorrhage\|brain inflammation | 884 |
| Coma | coma\|comas\|induced coma | 881 |
| Alopecia | hair loss\|hair fall\|losing hair | 792 |
| Sleepy | sleepiness\|sleeping\|sleepy | 752 |
| Feel a tickle | feel a tickle | 743 |
| Flu | flu - like symptoms\|flu like\|flu like symptoms\|flu symptoms | 734 |
| Heart failure | congestive heart failure\|damaged heart\|heart and lung damage\|heart attacks\|heart damage\|heart failure\|heart failures\|heart muscle damage | 690 |
| Sweating | sweating\|sweats\|night sweats\|sweat\|sweaty\|cold sweats | 665 |
| Respiratory failure | respiratory failure\|acute respiratory failure | 628 |
| Renal failure | acute renal failure\|renal failure | 626 |
| Circulatory failure | circulatory failure | 609 |
| Stomach ache | stomach pain\|stomach issues\|upset stomach\|stomach ache\|stomach problems | 589 |
| Immunosuppressed | autoimmune issues\|autoimmune reactions\|immune deficiency\|immune dysfunction\|immune suppression\|immunocompromised\|immunodeficiency\|immunosuppressed\|low immunity\|weak immunity | 571 |
| Syncope | faint\|fainting\|fainted\|vertigo | 562 |
| Impotence | erectile disfunction\|erectile dysfunction\|erection\|erections | 484 |
| Wheezing | wheezing\|wheeze\|wheezy | 461 |
| Swelling | brain swelling\|face swelled up\|facial swelling\|lung swelling\|swelling | 377 |
| Panic | panic\|panic attack\|panic attacks | 374 |
| Nasal congestion | stuffy\|nasal congestion\|stuffed up | 367 |
| Delirium | delirium | 303 |
| Nerve pain | nerve pain\|nerve damage | 302 |
| Loss of appetite | no appetite\|loss of appetite | 287 |

### Fig. A.1 Model construction

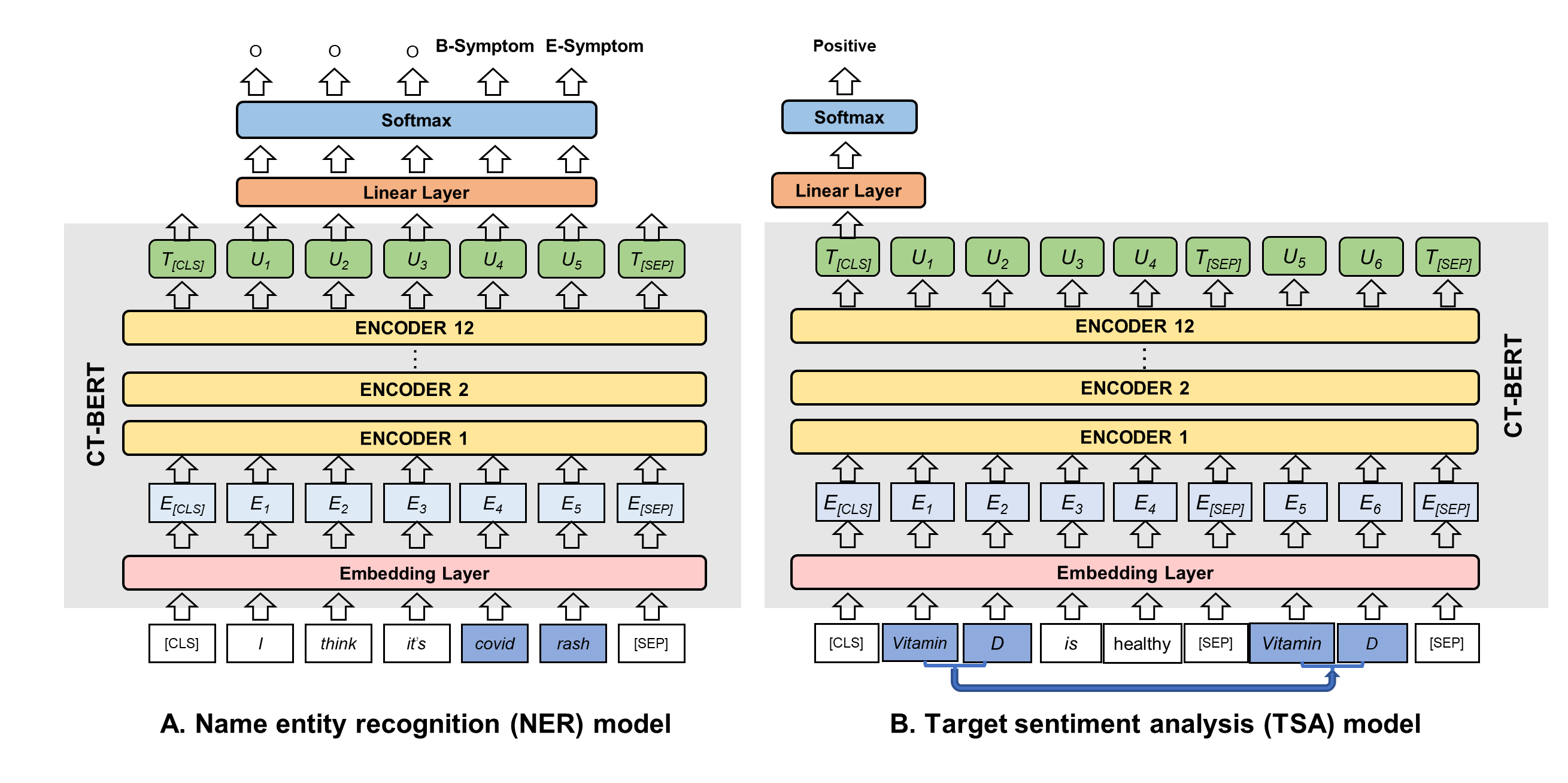

### Fig. A.2 Details of the text clustering preprocess

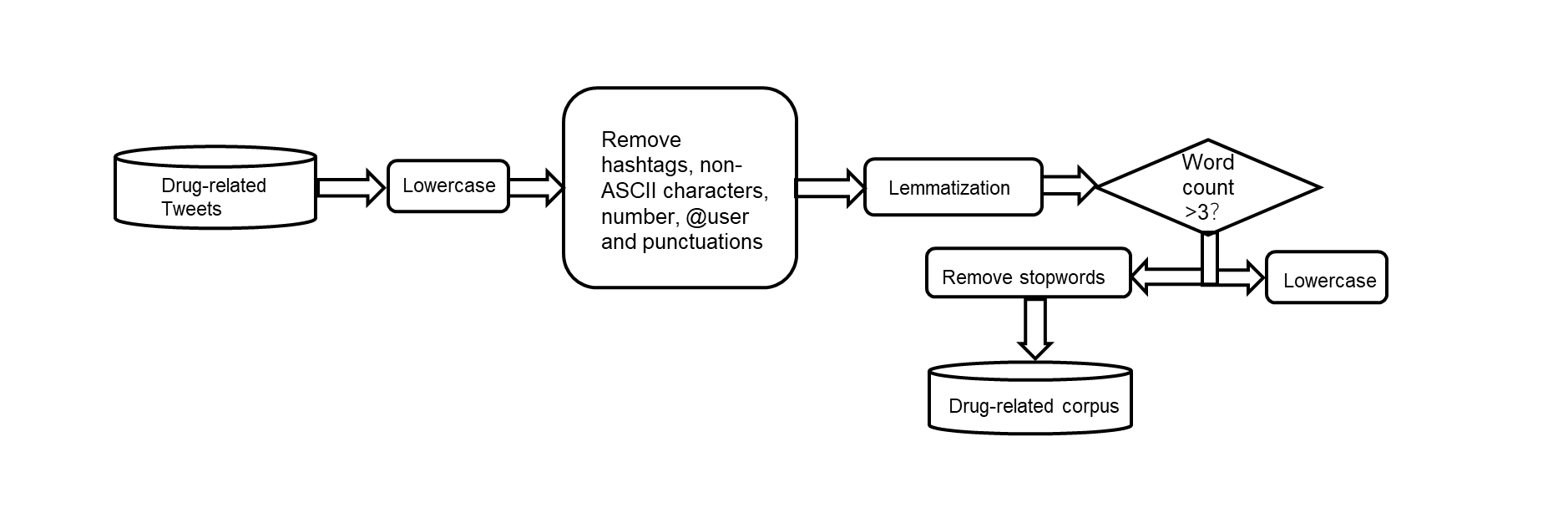

### Fig. A.3 Coherence score and perplexity of each topic

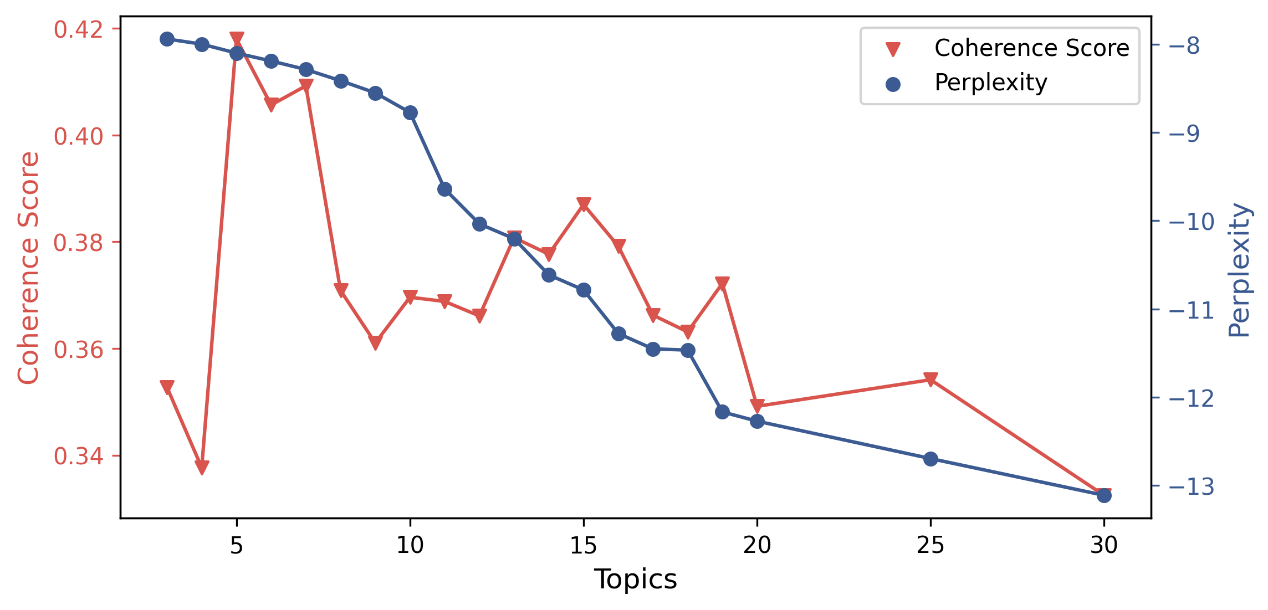

A lower degree of perplexity indicates that the model is less uncertain about which topic an article belongs to, and the better the clustering effect. Higher coherence scores indicate better interpretability of topics, meaning that the trained topics are more meaningful and semantically coherent. In our study, we chose the model with 15 topics when considering the lower perplexity, the higher coherence score, and the topic number.

**Fig. A.4 Top ten associations with five popular drugs**

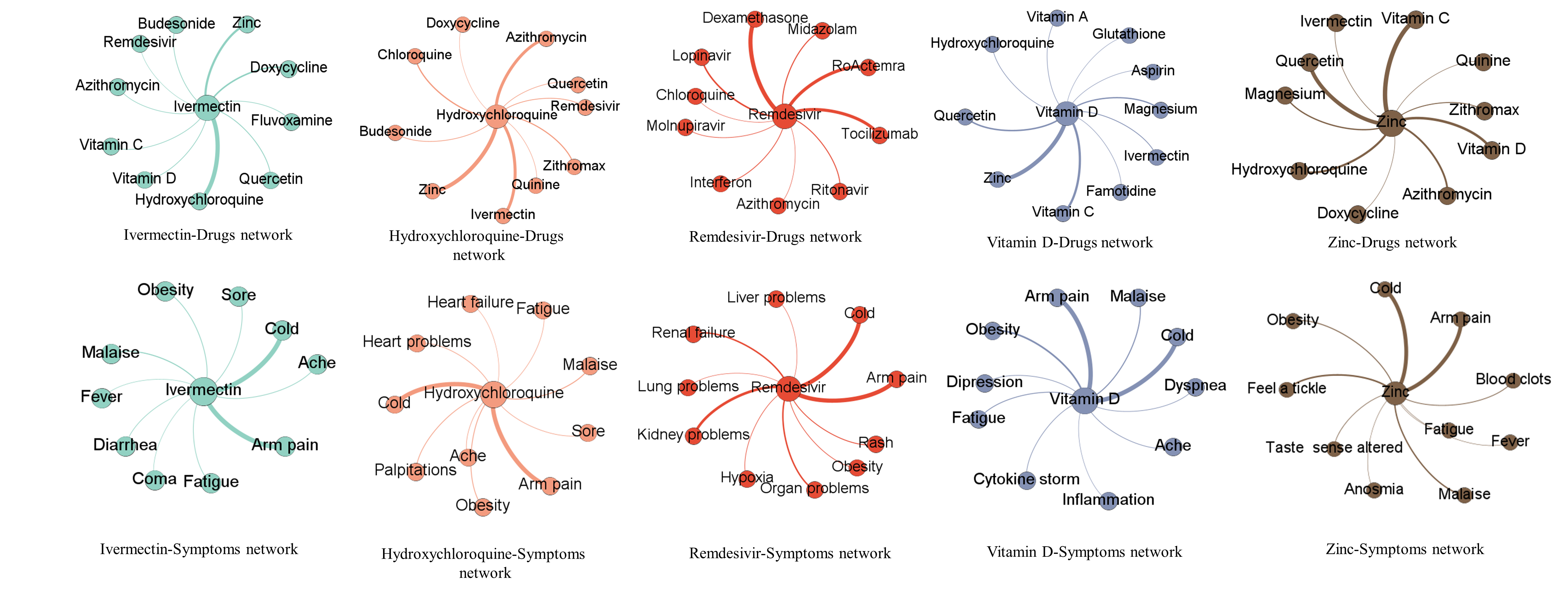

1. the Anatomical Therapeutic Chemical Classification System (ATC) from WHO, available at https://www.whocc.no/atc_ddd_index/. In situations where multiple classifications existed in one drug, the most widely used classification was selected.
2. Different representations of the same drug
